# Artificial Intelligence Enabled Prediction of Heart Failure Risk from Single-lead Electrocardiograms

**DOI:** 10.1101/2024.05.27.24307952

**Authors:** Lovedeep S Dhingra, Arya Aminorroaya, Aline F Pedroso, Akshay Khunte, Veer Sangha, Daniel McIntyre, Clara K Chow, Folkert W Asselbergs, Luisa CC Brant, Sandhi M Barreto, Antonio Luiz P Ribeiro, Harlan M Krumholz, Evangelos K Oikonomou, Rohan Khera

**Author notes:** Correspondence to: Rohan Khera, MD, MS, 195 Church Street, 6^th^ Floor, New Haven, CT 06510; @rohan_khera. Contributed equally as co-first authors (Lovedeep S Dhingra, Arya Aminorroaya).

## Abstract

**Importance:** Despite the availability of disease-modifying therapies, scalable strategies for heart failure (HF) risk stratification remain elusive. Portable devices capable of recording single-lead electrocardiograms (ECGs) can enable large-scale community-based risk assessment.

**Objective:** To evaluate an artificial intelligence (AI) algorithm to predict HF risk from noisy single-lead ECGs.

**Design:** Multicohort study.

**Setting:** Retrospective cohort of individuals with outpatient ECGs in the integrated Yale New Haven Health System (YNHHS) and prospective population-based cohorts of UK Biobank (UKB) and Brazilian Longitudinal Study of Adult Health (ELSA-Brasil).

**Participants:** Individuals without HF at baseline.

**Exposures:** AI-ECG-defined risk of left ventricular systolic dysfunction (LVSD).

**Main Outcomes and Measures:** Among individuals with ECGs, we isolated lead I ECGs and deployed a noise-adapted AI-ECG model trained to identify LVSD. We evaluated the association of the model probability with new-onset HF, defined as the first HF hospitalization. We compared the discrimination of AI-ECG against two risk scores for new-onset HF (PCP-HF and PREVENT equations) using Harrel’s C-statistic, integrated discrimination improvement (IDI), and net reclassification improvement (NRI).

**Results:** There were 192,667 YNHHS patients (age 56 years [IQR, 41-69], 112,082 women [58%]), 42,141 UKB participants (65 years [59-71], 21,795 women [52%]), and 13,454 ELSA-Brasil participants (56 years [41-69], 7,348 women [55%]) with baseline ECGs. A total of 3,697 developed HF in YNHHS over 4.6 years (2.8-6.6), 46 in UKB over 3.1 years (2.1-4.5), and 31 in ELSA-Brasil over 4.2 years (3.7-4.5). A positive AI-ECG screen was associated with a 3- to 7-fold higher risk for HF, and each 0.1 increment in the model probability portended a 27-65% higher hazard across cohorts, independent of age, sex, comorbidities, and competing risk of death. AI-ECG’s discrimination for new-onset HF was 0.725 in YNHHS, 0.792 in UKB, and 0.833 in ELSA-Brasil. Across cohorts, incorporating AI-ECG predictions in addition to PCP-HF and PREVENT equations resulted in improved Harrel’s C-statistic (Δ_PCP-HF_=0.112-0.114; Δ_PREVENT_=0.080-0.101). AI-ECG had IDI of 0.094-0.238 and 0.090-0.192, and NRI of 15.8%-48.8% and 12.8%-36.3%, vs. PCP-HF and PREVENT, respectively.

**Conclusions and Relevance:** Across multinational cohorts, a noise-adapted AI model defined HF risk using lead I ECGs, suggesting a potential portable and wearable device-based HF risk-stratification strategy.

**KEY POINTS:** *Question:* Can single-lead electrocardiograms (ECG) predict heart failure (HF) risk?

*Findings:* We evaluated a noise-adapted artificial intelligence (AI) algorithm for single-lead ECGs across multinational cohorts, spanning a diverse US health-system and community-based cohorts in the UK and Brazil. A positive AI-ECG screen was associated with 3- to 7-fold higher HF risk, independent of age, sex, and comorbidities. The AI model achieved incremental discrimination and improved reclassification over two established clinical risk scores for HF prediction.

*Meaning:* A noise-adapted AI model for single-lead ECG predicted the risk of new-onset HF, representing a scalable HF risk-stratification strategy for portable and wearable devices.

## BACKGROUND

Accessible strategies for risk stratification for heart failure (HF) remain elusive despite the availability of evidence-based therapies that can effectively modify the disease trajectory.^1,2^ Clinical scores to predict HF risk, such as the pooled cohort equations to prevent HF (PCP-HF), the predicting risk of cardiovascular disease events (PREVENT) equations, and the Health ABC score,^3–5^ require extensive clinical evaluation, including detailed history and physical exam, a 12-lead electrocardiogram (ECG), and other specialized testing.^3–9^ These complex inputs limit their use, systematically excluding those without healthcare access and preventing deployment for community-based screening.^8–10^ Similarly, serum-based biomarkers such as N-terminal pro–B-type natriuretic peptide (NT-proBNP) and high-sensitivity cardiac troponin, which portend a higher risk of HF when elevated, are limited by the need for blood draws and sample storage, and frequent inaccessibility at the point-of-contact.^11–16^ Thus, there is an unmet need for a simple and efficient strategy for identifying those at high risk for developing HF in the community.

Given their increasing utility and ubiquity, portable and wearable devices capable of recording single-lead ECG tracings have been proposed as a platform for cardiovascular monitoring and screening.^17–20^ Further, artificial intelligence (AI)-enhanced interpretation of ECGs (AI-ECG) has been shown to detect hidden cardiovascular disease signatures from single-lead ECGs, highlighting the potential of these devices for improving cardiovascular health.^21–25^ However, these portable ECGs are prone to noise introduction during acquisition, which can limit the AI model performance unless specialized measures are taken to ensure they are resilient to such noise.^21,26^ Recently, we reported a novel approach for single-lead ECGs that incorporates random noising during model development, enabling consistent diagnostic performance across varying levels of real-world noises.^21^ This initial deployment of our approach focused on developing a model that detects reduced left ventricular ejection fraction (LVEF) on single-lead ECG based on information from a concurrent echocardiogram, with the potential application of identifying subclinical left ventricular systolic dysfunction (LVSD). Recent studies also suggest that the AI-ECG signature for LVSD identifies other subtle markers of LV dysfunction, including abnormal LV strain and diastolic function, especially among those with a positive screen but preserved LVEF.^27^ Of note, each of these is associated with elevated HF risk.^28^

Given the increasing accessibility of single-lead ECGs, we tested the hypothesis that an AI model developed to detect the cross-sectional signature of LVSD from single-lead ECG tracings can predict future HF risk. We longitudinally evaluated our approach in individuals undergoing outpatient ECGs within a diverse US health system and two large population-based cohorts in the UK and Brazil.

## METHODS

### Data Sources

We included three large cohorts spanning different countries and settings who had undergone an ECG: (i) individuals seeking outpatient care in the Yale New Haven Health System (YNHHS), a large healthcare system in the Northeastern US, including five independent hospitals and an outpatient medical network, (ii) participants in the UK Biobank, a nationwide UK-based cohort study, and (iii) participants in the Brazilian Longitudinal Study of Adult Health (ELSA-Brasil), the largest community-based cohort study from Brazil. While YNHHS included testing and follow-up as a part of routine clinical care in an integrated health system, participants in UKB and ELSA-Brasil had detailed protocolized evaluation at baseline and comprehensive longitudinal follow-up (**eMethods**).

### Study Population

In YNHHS, to approximate a cohort resembling a screening setting, we identified patients undergoing a 12-lead ECG in an outpatient health encounter during 2014-2023 without HF before the ECG. To account for the ECGs potentially being obtained as a part of a workup for HF, we included a 1-year blanking period from the first recorded encounter in the electronic health records (EHR) to identify those with prevalent HF (**eMethods; eFigure 1**). In YNHHS, 255,604 individuals had at least one outpatient ECG after the blanking period. We excluded 47,720 patients in the original model development population and 11,954 patients with prevalent HF. To further focus the cohort on those without established precursors of HF, we excluded 1,590 patients with LV dysfunction (LVEF under 50% or moderate/severe LV diastolic dysfunction) and 1,673 patients with an NT-proBNP of >300 pg/mL before the index ECG (**eFigure 2**).

To avoid selection bias in UKB and ELSA-Brasil, we identified all participants who received an ECG as part of the study protocol. In UKB, we included 42,366 participants who had undergone a 12-lead ECG during their imaging study visit in 2014-2020. We used the linkage with the EHR from the UK National Health Service to exclude 225 participants who had been hospitalized with a principal or secondary discharge diagnosis of HF before the ECG. In ELSA-Brasil, we included 13,739 participants who had undergone a 12-lead ECG during 2008-2010, excluding those with an HF diagnosis (N=227) or with an LVEF under 50% (N=58) on their baseline echocardiogram (**eFigure 2**).

### Study Outcomes

We defined the study outcome as new-onset HF characterized by HF hospitalizations. In YNHHS, this was defined as a hospitalization with an International Classification of Disease Tenth Revision – Clinical Modification (ICD-10-CM) code for HF as the principal discharge diagnosis (**eTable 1**). This approach was guided by the over 95% specificity of HF diagnosis codes, especially as the principal discharge diagnosis, for a clinical diagnosis of HF.^29^ Similarly, in UKB, we used the linked EHR to identify hospitalizations with HF as the principal diagnosis code. In ELSA-Brasil, HF was identified by in-person interview or telephonic surveillance for all hospitalizations, followed by independent medical record review and adjudication of HF hospitalizations by two cardiologists (**eMethods**).^30^

We further evaluated the association of AI-ECG probabilities with alternate definitions of new-onset HF, and composite outcomes, including (i) any hospitalization with a principal or secondary HF diagnosis code, (ii) a subsequent echocardiogram with LVEF under 50%, and (iii) a composite outcome of HF or all-cause death (**eMethods**). To evaluate the specificity of AI-ECG-defined HF risk, we examined the risk of other cardiovascular conditions, including acute myocardial infarction (AMI), stroke hospitalizations, and all-cause mortality (**eTable 1**). A composite outcome of major adverse cardiovascular events (MACE) was defined as death or hospitalization for HF, AMI, or stroke.

### Study Exposure

We defined the study exposure as the output probability of an AI-ECG model trained to detect concurrent LVSD on lead I of a 12-lead ECG, representing the lead commonly captured by portable ECG devices.^21^ This was developed at the Yale New Haven Hospital (YNHH) using a novel approach of augmenting training data with random Gaussian noise (**eMethods**). The model achieved excellent discrimination (area under the receiver operating characteristic curve of 0.899 [95% CI, 0.889-0.909]) for detecting concurrent LVSD in the YNHH held-out test set and performed consistently across clinical and population-based external validation cohorts (**eTable 2; eFigure 3**).

We deployed this established model without further development to lead I ECG signals across included individuals to obtain the LVSD probability, representing a continuous HF risk score. We defined a positive AI-ECG screen as an output probability greater than 0.08, representing the model threshold for 90% sensitivity for detecting LVSD during internal validation.^21^

### Study Comparator

We compared the performance of the AI-ECG algorithm with established risk scores for predicting HF risk, including the PCP-HF and the PREVENT equations. The PCP-HF score was developed and validated in 7 community-based cohorts.^4^ It uses 12 input features, including demographics (age, sex, race), physical exam-based features (smoking status, BMI, systolic blood pressure), laboratory measurements (total cholesterol, high-density lipoprotein cholesterol, fasting blood glucose), medication history (use of antihypertensive and antihyperglycemic medications), and electrocardiographically-defined QRS duration. The PREVENT equations were recently developed using data from over 3.2 million individuals and were validated in 21 datasets.^5,33^ The PREVENT equations for HF risk prediction employ 8 inputs, entailing demographics (age, sex), medical history (type 2 diabetes mellitus), physical exam-based features (smoking status, BMI, systolic blood pressure), laboratory measurements (estimated glomerular filtration rate), and medication history (antihypertensive medication use). Across cohorts, these features were determined using the EHR and/or study visits (**eMethods**).^34–37^

### Statistical Analysis

We used age-, sex-, and comorbidity-adjusted Cox proportional hazard models with time-to-first HF event as the dependent variable and the AI-ECG-based screen results (positive/negative) or continuous model probability as the independent variable to evaluate the association of the model output with HF risk. Multi-outcome Fine-Gray subdistribution hazard models were used to account for the competing risk of death.^38^

The incremental discrimination of AI-ECG over PCP-HF and PREVENT equations for predicting time-to-HF hospitalization was reported as the difference in Harrel’s C-statistics and 95% confidence interval (CI) using a one-shot nonparametric approach.^39^ We calculated integrated discrimination improvement (IDI), and categorical and continuous time-to-event net reclassification improvement (NRI).^40^ We further compared the net benefit of the AI-ECG model with the PCP-HF and PREVENT equations across all probability thresholds (**eMethods)**.^41^

## RESULTS

### Study Population

From YNHHS, we included 192,667 individuals with a median age of 56 years (IQR, 41-69), comprising 111,181 (57.7%) women, 117,857 (61.2%) non-Hispanic White, 30,623 (15.9%) non-Hispanic Black, and 33,256 (17.3%) Hispanic individuals. Over a median 4.6-year follow-up (IQR, 2.8-6.6), 3,697 (1.9%) had an HF hospitalization, 7,514 (3.9%) had an HF hospitalization or an LVEF below 50% on subsequent echocardiogram, and 10,381 (5.4%) died (**Table 1, eTable 3**).

**Figure 1.**
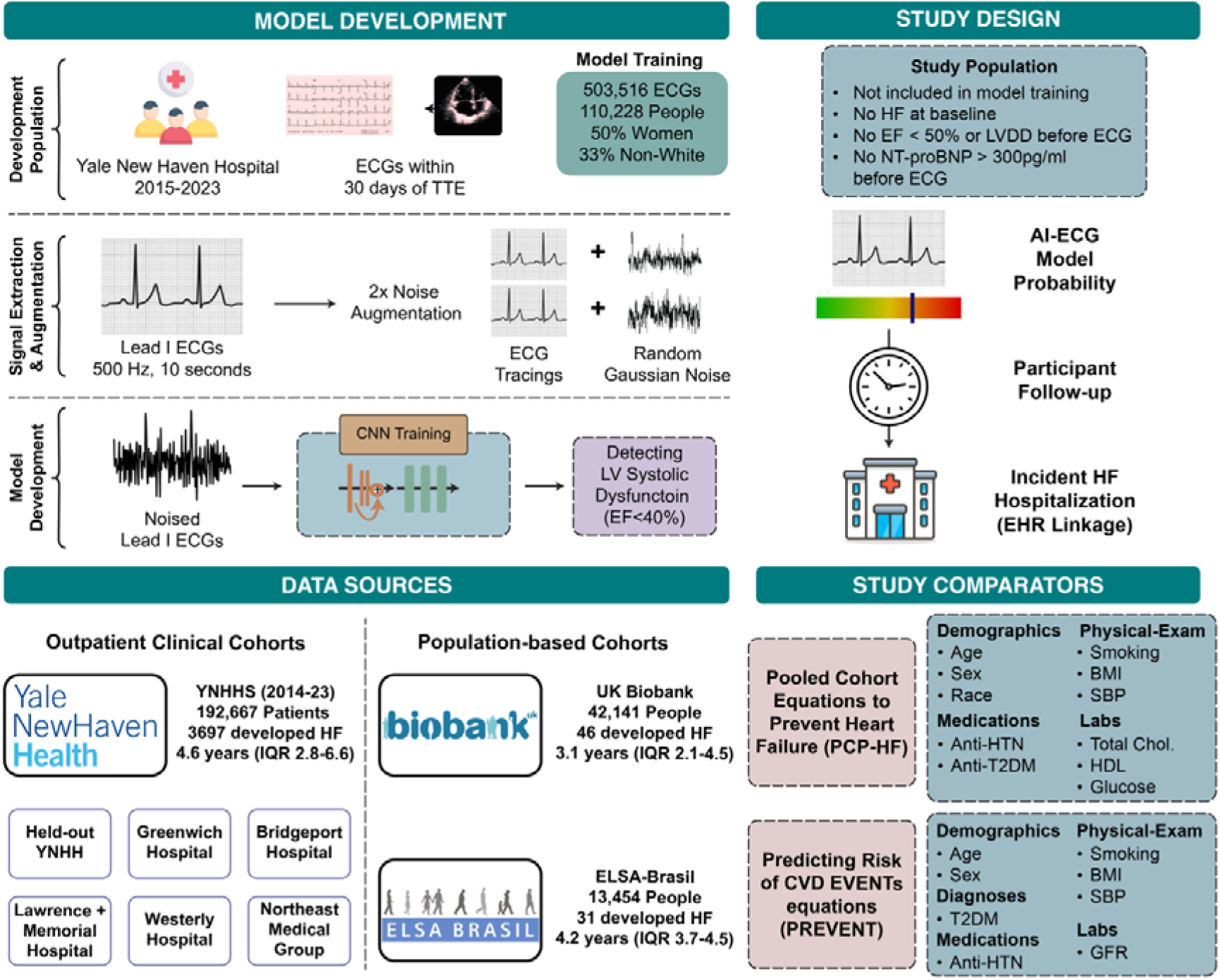
Study Overview. Abbreviations: BMI, Body Mass Index; BP; Blood Pressure; CNN, Convolutional Neural Network; ECG, Electrocardiogram; EF, Ejection Fraction; EHR, Electronic Health Record; ELSA-Brasil, Brazilian Longitudinal Study of Adult Health; HDL, High-density Lipoprotein Cholesterol; HF, Heart Failure; LV, Left Ventricle; YNH, Yale New Haven Hospital; YNHHS, Yale New Haven Health System.

**Table 1.**
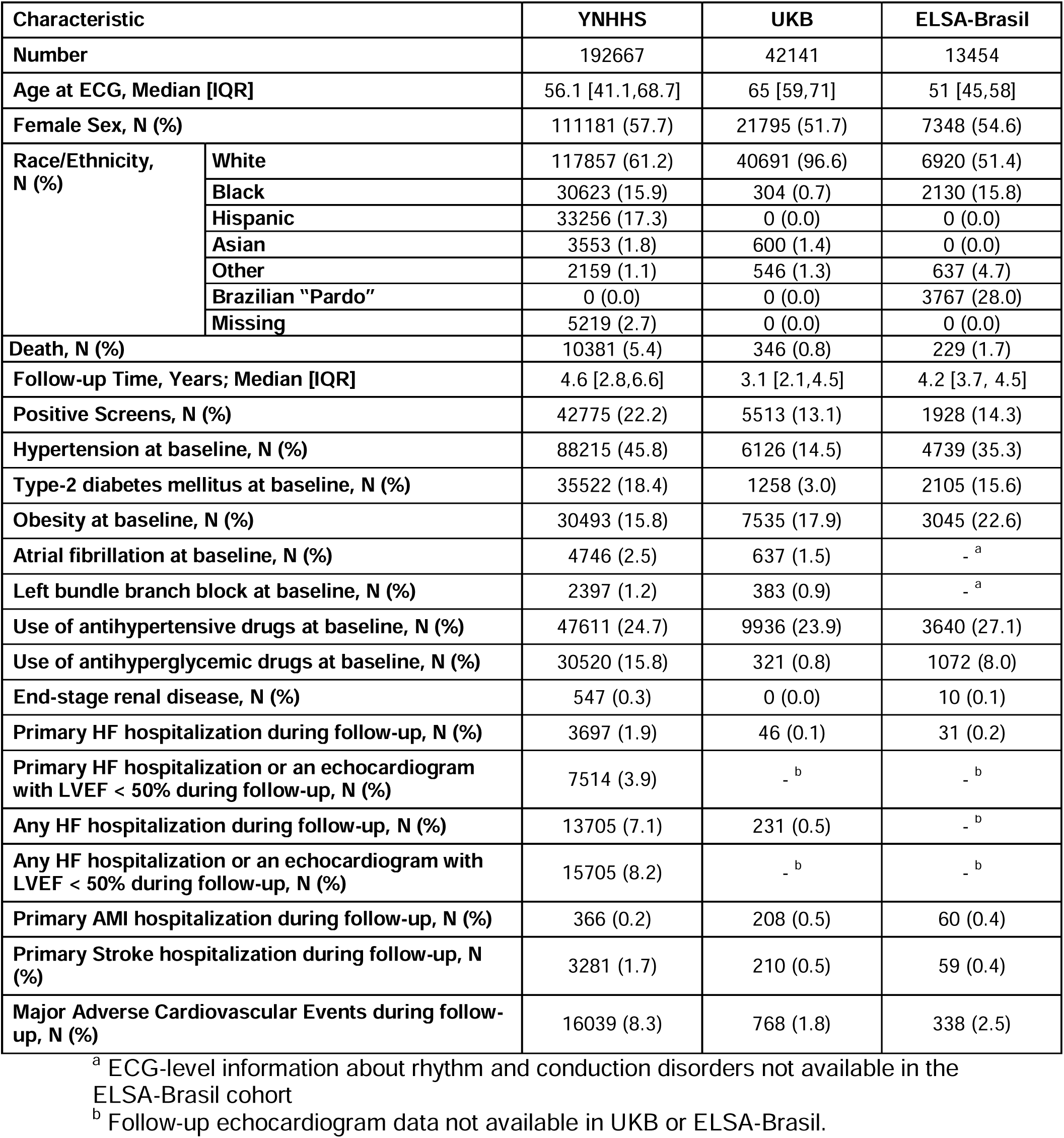
Population Characteristics of the Study Cohorts. Abbreviations: AMI, acute myocardial infarction; ECG, Electrocardiogram; ELSA-Brasil, Brazilian Longitudinal Study of Adult Health; HF, heart failure; IQR, Interquartile Range; LVEF, Left Ventricular Ejection Fraction; UKB, UK Biobank; YNHHS, Yale New Haven Health System.

The 42,141 UKB participants had a median age of 65 years (IQR, 59-71), including 21,795 (51.7%) women, with 40,691 (96.6%) identifying as White and 304 (0.7%) as Black. Over a median follow-up of 3.1 years (IQR, 2.1-4.5), 46 (0.1%) had an HF hospitalization, and 346 (0.8%) died (**Table 1**).

From ELSA-Brasil, the 13,454 participants had a median age of 51 years (IQR, 45-58), comprising 7,348 (54.6%) women, 6,920 (51.4%) adults identifying as White, 2,130 (15.8%) as Black, and 3,767 (28.0%) as “Pardo”. Over a median of 4.2 years (IQR, 3.7-4.5), 31 (0.2%) people developed HF, and 229 (1.7%) died.

### Risk Stratification for New-Onset HF

In YNHHS, 42,775 (22.2%) patients screened positive on the AI model applied to the baseline single-lead ECG signal. A positive screen was associated with over 5-fold higher risk of developing HF (HR 5.05 [95% CI, 4.73-5.39]; **Table 2**). After accounting for differences in age and sex, a positive AI-ECG screen was associated with a 3.3-fold higher risk of HF compared with a negative screen (adjusted HR [aHR], 3.31 [95% CI, 3.10-3.54]). The association remained statistically significant after accounting for differences in HF risk factors of prior ischemic heart disease, hypertension, type 2 diabetes, and obesity (aHR 2.81 [95% CI, 2.63-3.01]) and after additionally accounting for the competing risk of death (aHR of 2.73 [95% CI, 2.55-2.93]). The association of a positive screen with an elevated risk of HF was noted across YNHHS sites (**eTable 4**), demographic subgroups (**eTable 5**), and different HF definitions (**eTables 6-7**).

**Table 2.** Model Performance for Predicting Heart Failure Risk Based on AI-ECG Probability. Abbreviations: ELSA-Brasil, Brazilian Longitudinal Study of Adult Health; IHD, Ischemic Heart Disease; HTN, hypertension; T2DM, type-2 diabetes mellitus; UKB, UK Biobank; YNHHS, Yale New Haven Health System.

| Model Type | Predictive Model Inputs | YNHHS |  | UKB |  | ELSA-Brasil |  |
| --- | --- | --- | --- | --- | --- | --- | --- |
|  |  | Positive Screen | Per 0.1 Increment | Positive Screen | Per 0.1 Increment | Positive Screen | Per 0.1 Increment |
| **Cox Proportional Hazard Model** | AI-ECG Probability | 5.05 (4.73-5.39) | 1.45 (1.44-1.47) | 7.52 (4.21-13.41) | 1.55 (1.40-1.71) | 11.11 (5.32-23.19) | 1.83 (1.64-2.05) |
| **Cox Proportional Hazard Model** | AI-ECG Probability + Age + Sex | 3.31 (3.10-3.54) | 1.32 (1.30-1.34) | 5.96 (3.32-10.68) | 1.52 (1.37-1.68) | 8.74 (4.13-18.48) | 1.75 (1.56-1.97) |
| **Cox Proportional Hazard Model** | AI-ECG Probability + Age + Sex + IHD + HTN + T2DM + Obesity | 2.81 (2.63-3.01) | 1.28 (1.26-1.30) | 5.02 (2.77-9.09) | 1.49 (1.33-1.66) | 7.71 (3.62-16.46) | 1.72 (1.52-1.93) |
| **Fine-Gray Subdistribution Hazard Model** | AI-ECG Probability + Age + Sex and accounting for competing risk of death | 3.22 (3.01-3.45) | 1.30 (1.29-1.32) | 5.91 (3.33-10.50) | 1.51 (1.38-1.66) | 8.67 (4.02-18.70) | 1.74 (1.55-1.96) |
| **Fine-Gray Subdistribution Hazard Model** | AI-ECG Probability + Age + Sex + IHD + HTN + T2DM + Obesity and accounting for competing risk of death | 2.73 (2.55-2.93) | 1.27 (1.25-1.28) | 4.99 (2.81-8.87) | 1.49 (1.36-1.63) | 6.53 (2.91-14.67) | 1.65 (1.46-1.87) |

In UKB, 5,513 (13.1%) participants screened positive with the AI-ECG model. A positive AI-ECG screen portended a 7.5-fold higher hazard for developing HF (HR 7.52 [95% CI, 4.21-13.41]). After accounting for age, sex, HF risk factors, and the competing risk of death, screen-positive participants had a 5-fold higher risk of HF (aHR 5.02 [95% CI, 2.77-9.09]; **Table 2**).

In the ELSA-Brasil cohort, 1,928 (14.3%) participants had a positive AI-ECG screen, with a 9-fold higher risk for HF (age- and sex-adjusted HR 8.74 [95% CI, 4.13-18.48]) compared with screen-negative participants. This association was consistent even after accounting for the comorbidities and the competing risk of death (aHR 7.71 [95% CI, 3.62-16.46]).

### Risk Across Model Probability Increments

Across the YNHHS network, each 0.1 increment in the model output probability portended a 28% higher hazard of developing HF, adjusted for age, sex, comorbidities, and accounting for the competing risk of death (aHR 1.28 [95% CI, 1.26-1.30]; **Table 2**). Higher model probabilities were progressively associated with higher HF risk across various probability bins, with consistent patterns across the individual hospitals and the outpatient medical network (**eFigure 4**, **eTables 4, 8-9**).

Across both UKB and ELSA-Brasil cohorts, a 0.1 increment in model probability was associated with 51% and 66% higher adjusted risk of HF (aHR 1.49 [95% CI, 1.36-1.63] and aHR 1.65 [95% CI, 1.46-1.87], respectively, **Table 2**), respectively.

### Comparison with PCP-HF and PREVENT Equations

The AI-ECG model had a discrimination based on Harrel’s C-statistic of 0.723 (95% CI, 0.701-0.744) in YNHHS, compared with 0.634 (95% CI, 0.612-0.656) for PCP-HF (p<0.001), and 0.668 (95% CI, 0.644-0.693) for PREVENT equations (p< 0.001; **Table 3**). In UKB and ELSA-Brasil, the AI-ECG model’s discrimination for HF was 0.792 (95% CI, 0.696-0.889) and 0.833 (95% CI, 0.720-0.946), respectively, which was not significantly different from the clinical risk scores (AI-ECG vs PCP-HF: p_UKB_=0.45; p_ELSA-Brasil_=0.87; AI-ECG vs PREVENT: p_UKB_=0.90; p_ELSA-Brasil_=0.92). Across cohorts, incorporating AI-ECG predictions in addition to PCP-HF and PREVENT equations resulted in improved Harrel’s C-statistic (Δ_PCP-HF_=0.112-0.114; Δ_PREVENT_=0.080-0.101), compared with the use of the clinical risk equations alone (**Table 3**).

**Table 3.**
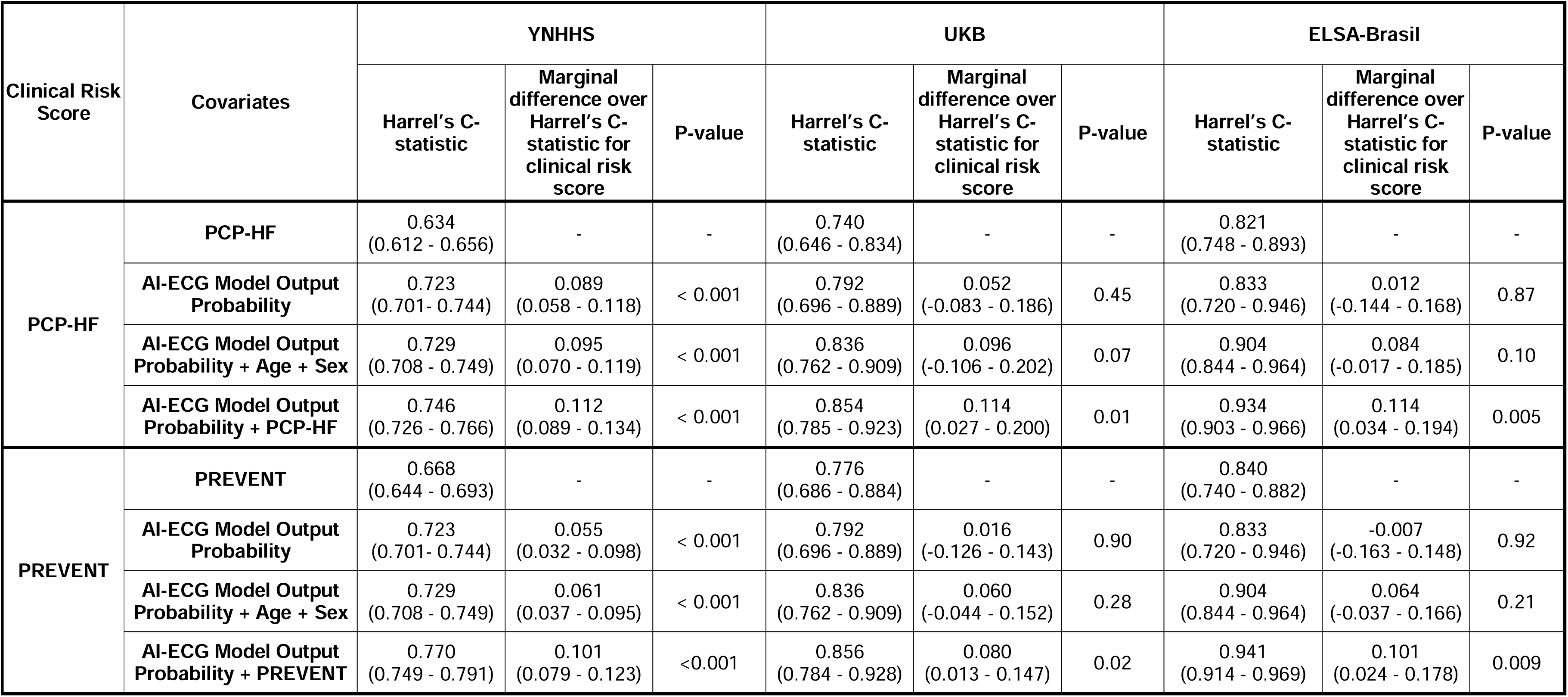
Comparison of Discrimination for AI-ECG Model Output Probability with Pooled Cohort Equations to Prevent Heart Failure and Predicting Risk of Cardiovascular Disease Events Equations for Predicting Incident Heart Failure. Abbreviations: ELSA-Brasil, Brazilian Longitudinal Study of Adult Health; PCP-HF, Pooled Cohort Equations to Prevent Heart Failure; PREVENT, Predicting Risk of Cardiovascular Disease Events; UKB, UK Biobank; YNHHS, Yale New Haven Health System.

| Clinical Risk Score | Covariates | YNHHS |  |  | UKB |  |  | ELSA-Brasil |  |  |
| --- | --- | --- | --- | --- | --- | --- | --- | --- | --- | --- |
|  |  | Harrel's C-statistic | Marginal difference over Harrel's C-statistic for clinical risk score | P-value | Harrel's C-statistic | Marginal difference over Harrel's C-statistic for clinical risk score | P-value | Harrel's C-statistic | Marginal difference over Harrel's C-statistic for clinical risk score | P-value |
| PCP-HF | PCP-HF | 0.634<br>(0.612 - 0.656) | - | - | 0.740<br>(0.646 - 0.834) | - | - | 0.821<br>(0.748 - 0.893) | - | - |
|  | AI-ECG Model Output Probability | 0.723<br>(0.701 - 0.744) | 0.089<br>(0.058 - 0.118) | < 0.001 | 0.792<br>(0.696 - 0.889) | 0.052<br>(-0.083 - 0.186) | 0.45 | 0.833<br>(0.720 - 0.946) | 0.012<br>(-0.144 - 0.168) | 0.87 |
|  | AI-ECG Model Output Probability + Age + Sex | 0.729<br>(0.708 - 0.749) | 0.095<br>(0.070 - 0.119) | < 0.001 | 0.836<br>(0.762 - 0.909) | 0.096<br>(-0.106 - 0.202) | 0.07 | 0.904<br>(0.844 - 0.964) | 0.084<br>(-0.017 - 0.185) | 0.10 |
|  | AI-ECG Model Output Probability + PCP-HF | 0.746<br>(0.726 - 0.766) | 0.112<br>(0.089 - 0.134) | < 0.001 | 0.854<br>(0.785 - 0.923) | 0.114<br>(0.027 - 0.200) | 0.01 | 0.934<br>(0.903 - 0.966) | 0.114<br>(0.034 - 0.194) | 0.005 |
| PREVENT | PREVENT | 0.668<br>(0.644 - 0.693) | - | - | 0.776<br>(0.686 - 0.884) | - | - | 0.840<br>(0.740 - 0.882) | - | - |
|  | AI-ECG Model Output Probability | 0.723<br>(0.701 - 0.744) | 0.055<br>(0.032 - 0.098) | < 0.001 | 0.792<br>(0.696 - 0.889) | 0.016<br>(-0.126 - 0.143) | 0.90 | 0.833<br>(0.720 - 0.946) | -0.007<br>(-0.163 - 0.148) | 0.92 |
|  | AI-ECG Model Output Probability + Age + Sex | 0.729<br>(0.708 - 0.749) | 0.061<br>(0.037 - 0.095) | < 0.001 | 0.836<br>(0.762 - 0.909) | 0.060<br>(-0.044 - 0.152) | 0.28 | 0.904<br>(0.844 - 0.964) | 0.064<br>(-0.037 - 0.166) | 0.21 |
|  | AI-ECG Model Output Probability + PREVENT | 0.770<br>(0.749 - 0.791) | 0.101<br>(0.079 - 0.123) | <0.001 | 0.856<br>(0.784 - 0.928) | 0.080<br>(0.013 - 0.147) | 0.02 | 0.941<br>(0.914 - 0.969) | 0.101<br>(0.024 - 0.178) | 0.009 |

Compared with the PCP-HF and PREVENT scores, the AI-ECG algorithm had a positive IDI across all study cohorts. The AI model was associated with a significant improvement in continuous NRI at YNHHS, but not in the UKB and ELSA-Brasil (**Table 4**). However, the AI model significantly improved categorical NRI across all data sources. This improvement in categorical NRI was driven by improved event NRI, reclassification of cases, while non-event NRI decreased (**eTable 10**). Despite the differential improvement in reclassifying cases and controls, the AI-ECG’s PPV was comparable with PCP-HF and PREVENT across sites (**eTable 11**). The AI-ECG model demonstrated consistent superior net benefit over PCP-HF across all probability thresholds in UKB and ELSA-Brasil. In YNHHS, the net benefit of AI-ECG was higher than that of PCP-HF and PREVENT equations in probability thresholds greater than 0.04, where the AI-ECG threshold is 0.08 (**eFigure 5**). A positive AI-ECG screen was an independent predictor of HF risk after accounting for PCP-HF score, with consistent patterns across racial groups (**eFigures 6-7**).

**Table 4.** Integrated Discrimination Improvement and Categorical and Continuous Time-to-Event Net Reclassification Index of AI-ECG Model Output Probability over Pooled Cohort Equations to Prevent Heart Failure and Predicting Risk of Cardiovascular Disease Events Equations for Heart Failure. Abbreviations: ELSA-Brasil, Brazilian Longitudinal Study of Adult Health; IDI, Integrated Discrimination Improvement; NRI, Net Reclassification Index; PCP-HF, Pooled Cohort Equations to Prevent Heart Failure; PREVENT, Predicting Risk of Cardiovascular Disease Events; UKB, UK Biobank; YNHHS, Yale New Haven Health System.

| Metric | YNHHS |  | UKB |  | ELSA-Brasil |  |
| --- | --- | --- | --- | --- | --- | --- |
|  | PCP-HF | PREVENT | PCP-HF | PREVENT | PCP-HF | PREVENT |
| **IDI** | 0.094 (0.075 to 0.114) | 0.090 (0.077 to 0.105) | 0.165 (0.069 to 0.265) | 0.153 (0.076 to 0.243) | 0.238 (0.107 to 0.372) | 0.192 (0.099 to 0.295) |
| **Categorical NRI** | 0.158 (0.102 to 0.218) | 0.128 (0.092 to 0.167) | 0.291 (0.092 to 0.494) | 0.330 (0.088 to 0.547) | 0.488 (0.188 to 0.741) | 0.363 (0.118 to 0.592) |
| **Continuous NRI** | 0.199 (0.116 to 0.283) | 0.214 (0.161 to 0.275) | 0.155 (-0.211 to 0.478) | 0.366 (0.001 to 0.701) | 0.077 (-0.238 to 0.324) | 0.122 (-0.204 to 0.455) |

### Non-HF Cardiovascular Outcome Prediction

In YNHHS, a positive AI-ECG screen was associated with a modestly elevated risk of stroke and MACE (age- and sex-adjusted HRs: stroke, 1.17; MACE, 1.77; **eTables 12-13**) compared with a 3-fold increase in HF risk. In UKB and ELSA-Brasil, a positive screen portended a 1.5- to 4-fold hazard of stroke, death, and MACE compared with a 6- to 9-fold increase in HF risk.

## DISCUSSION

Across clinically and geographically distinct cohorts, a noise-adapted AI model, trained to detect cross-sectional LVSD from only a lead I ECG, predicted the risk of future HF among individuals seeking outpatient care and community-dwelling adults. Individuals with a positive AI-ECG screen had a 3- to 7-fold higher risk of developing HF compared with those with a negative screen, independent of known demographic and clinical risk factors. Higher AI-ECG probabilities were progressively associated with a higher risk of HF, with each 10% increment portending a 27-65% higher risk-adjusted hazard for HF across all cohorts. Further, the AI-ECG model demonstrated incremental discrimination, improved reclassification, and superior net benefit over the PCP-HF score. Therefore, our AI-based approach presents ideal characteristics for use as a non-invasive digital biomarker for elevated HF risk using a single-lead ECG.

Applications of deep learning for ECGs have demonstrated the ability to identify subtle signatures of structural heart disorders previously considered electrically silent,^42–51^ with applications extending to detecting LVSD from single-lead tracings.^21,52–55^ Further, the US Food and Drug Administration recently cleared an AI tool using electronic stethoscope-based single-lead ECGs for cross-sectional LVSD detection.^56^ Our study demonstrates that a noise-adapted AI-ECG model can predict new-onset HF risk using single-lead ECG tracings. Given the increasing accessibility of portable and wearable devices capable of acquiring ECG signals outside a clinical setting,^20,57,58^ this approach can potentially be applied widely to identify individuals at a high risk of HF.^58^ While the ECGs acquired with these devices are often distorted by positioning and movement of electrodes or artifacts due to skeletal muscle contraction during acquisition,^26,59^ our unique noise-adapted training approach can enable reliable inference from these noisy ECGs.^56^

In this study, we opted for a definition of HF based on the principal discharge diagnosis code, a criterion with high specificity.^29^ Nonetheless, the association of a positive screen with elevated HF risk was consistent across several sensitivity analyses defining the condition differently in YNHHS and UKB, and in ELSA-Brasil where the outcomes were explicitly adjudicated. The robust performance across clinically and demographically distinct cohorts indicates that the model captures a predictive HF signature independent of site-specific coding practices.^61–63^ Moreover, the dose-dependent association of higher AI-ECG scores with progressively elevated HF risk enables graded risk stratification and risk-informed management. Notably, while a positive screen was also associated with a modestly elevated risk of other cardiovascular outcomes, including MACE, the predictive signature was more specific for HF.

Our study has important implications for defining HF risk. While several clinical risk score- and serum biomarker-based strategies have been proposed to identify those at high risk, these strategies often require extensive clinical evaluation and blood testing.^61,62^ This limits their scope to patients with established access to healthcare services.^9,16,64–66^ In contrast, our AI-based approach using single-lead ECG tracings may offer a means for HF risk stratification outside clinical settings. Notably, the model demonstrated positive IDI, improved reclassification, and greater net benefit compared with PCP-HF and PREVENT scores. Across all sites, the model also had incremental discrimination over the use of the clinical risk scores alone. While AI-ECG had comparable discrimination and did not improve continuous NRI over PCP-HF and PREVENT scores in the UKB and ELSA-Brasil, it showed a consistent improvement in categorical NRI. The categorical NRI is relevant for clinical decision-making, as it considers thresholds for guiding treatment and follow-up.^60^ Furthermore, the model’s PPV was comparable to PCP-HF and PREVENT, suggesting that an AI-ECG-based strategy for screening will not lead to unnecessary additional testing.

The ability to use a single portable device to record ECGs for multiple individuals could support the design of efficient community-based screening programs.^67,68^ Successful health promotion strategies, such as targeted hypertension management in barbershops and cancer screening in churches across the US,^69,70^ can be adapted to promote HF screening, especially among those traditionally less likely to seek preventive medical care.^64^ The ease of use and the brief time for ECG acquisition with these devices can enable a non-laboratory-based strategy, potentially suitable for integration into national-level non-communicable disease screening programs globally, especially in low- and middle-income countries.^67,68,71^ This scalability and potential community health benefits necessitate prospective clinical and cost-effectiveness assessments for AI-based HF risk stratification.

Our study has certain limitations. First, waveforms extracted from the lead I of clinical ECGs may not be identical to those from portable devices. While our noise-augmentation approach previously demonstrated sustained performance on ECGs with real-world noises,^70,71^ prospective validation of the model on portable- and wearable-acquired ECGs is necessary before deployment for community HF screening. This includes evaluating device types, acquisition methods, preprocessing techniques, and handling of ECG segments of longer durations. Second, despite YNHHS’s wide geographic coverage, out-of-hospital clinical outcomes may not have been captured, thereby representing a lower HF risk compared with the protocolized follow-up in UKB and ELSA-Brasil. Moreover, while we included only ECGs performed in an outpatient setting, the patients who underwent ECG testing were clinically selected, indicating an unmeasured potential risk profile of those who underwent a clinical ECG but had a negative AI-ECG screen. However, the controls in this setting all underwent ECG screens as well. Third, the number of HF outcome events was low in the UKB and ELSA-Brasil. However, the HF hospitalization rates were comparable to other population-based cohorts,^72,73^ and the outcome capture and adjudication in UKB and ELSA-Brasil have been extensively validated.^31,74–78^ Fourth, given the lack of NT-proBNP assessments in UKB and ELSA-Brasil, we could not evaluate NT-proBNP as a comparator in this study. In YNHHS, the use of NT-proBNP could incorporate substantial selection bias, since it is typically ordered for evaluation of cardiopulmonary symptoms and rarely for primary prevention. Nevertheless, a future head-to-head assessment of AI-ECG and NT-proBNP as predictors for HF is warranted. Further, while performed an analysis that excluded individuals with elevated pre-ECG NT-proBNP levels in YNHHS, the lack of NT-proBNP testing precluded this analysis in UKB and ELSA-Brasil. Finally, while the AI-ECG approach identifies individuals at elevated HF risk, it is not clear if this risk is modifiable. Nonetheless, a robust screening strategy can enable targeted management of known HF risk factors.

## CONCLUSION

Across clinically and geographically distinct cohorts, we used a noise-resilient AI model with a lead I ECG tracing as the sole input to define the risk of future HF, with value over conventional risk scores in both performance and ease of deployment. With the increasing availability of single-lead ECGs on portable and wearable devices, this AI-ECG-based non-invasive digital biomarker can enable scalable stratification of HF risk across communities.

## Supporting information

Supplemental Content

## Data Availability

The analyzed de-identified data are available for the UK Biobank cohort from the UK Biobank's Access Management System, and for the ELSA-Brasil cohort upon a reasonable request to the corresponding author. Individual-level data for the YNHHS cohort cannot be made available due to HIPAA regulations enforced by the Yale IRB. Additional supporting information (statistical/analytic code) is available upon request to the corresponding author.

## Author Contributions

Drs Dhingra, Aminorroaya, and Khera had full access to all of the data in the study and take responsibility for the integrity of the data and the accuracy of the data analysis. All authors approved the final version for submission.

*Study concept and design*: Dhingra, Lovedeep S; Aminorroaya, Arya; Oikonomou, Evangelos K; Khera, Rohan.

*Acquisition, analysis, or interpretation of data*: Dhingra, Lovedeep S; Aminorroaya, Arya; Pedroso Camargos, Aline; Khunte, Akshay; Sangha, Veer; McIntyre, Daniel; Chow, Clara K; Asselbergs, Folkert W; Brant, Luisa CC; M Barreto, Sandhi; Ribeiro, Antonio Luiz P; Krumholz, Harlan M; Oikonomou, Evangelos K; Khera, Rohan *Drafting of the manuscript*: Dhingra, Lovedeep S; Aminorroaya, Arya; Pedroso Camargos, Aline.

*Critical revision of the manuscript for important intellectual content*: Khunte, Akshay; Sangha, Veer; McIntyre, Daniel; Chow, Clara K; Asselbergs, Folkert W; Brant, Luisa CC; M Barreto, Sandhi; Ribeiro, Antonio Luiz P; Krumholz, Harlan M; Oikonomou, Evangelos K; Khera, Rohan.

*Statistical analysis*: Dhingra, Lovedeep S; Aminorroaya, Arya.

*Obtained funding*: Khera, Rohan.

*Administrative, technical, or material support*: Aminorroaya, Arya; Dhingra, Lovedeep S; Pedroso Camargos, Aline; Khera, Rohan.

*Study supervision*: Khera, Rohan.

## Disclaimer

The views expressed in this article are those of the authors and not necessarily any funders.

## Data Sharing Statement

The data from the Yale New Haven Health System represent protected health information. To protect patient privacy, the Yale Institutional Review Board does not allow the sharing of these data. Data from the UK Biobank and the Brazilian Longitudinal Study of Adult Health are available for research to licensed users. The code for cohort creation and statistical analyses is publicly available at https://github.com/CarDS-Yale/AI-ECG-HF-Pred.

