## Supplemental Content for "Artificial Intelligence Enabled Prediction of Heart Failure Risk from Single-lead Electrocardiograms"

### **eMethods**

**Data Sources**

The Yale New Haven Health System (YNHHS) is the largest referral center in southern New England and serves a diverse patient population. The YNHHS includes five hospitals, Yale New Haven Hospital, Bridgeport Hospital, Greenwich Hospital, Lawrence and Memorial Hospital, and Westerly Hospital, and a large network of community outpatient clinics, the Northeast Medical Group. The electronic health records (EHR) data was acquired during patient care at YNHHS using Epic and was extracted from the Clarity database.^1,2^

UK Biobank (UKB) is a prospective cohort of 502,468 community-dwelling adults aged 40-69 years recruited during 2006-2010.^3^ A group of these participants accepted to participate in the third or fourth UKB study visit during which the participants underwent 12-lead electrocardiograms (ECGs) in 2014-2021. The UKB dataset is linked with the national EHR from the UK National Health Service predating UKB enrollment, enabling access to EHR diagnosis codes.^4,5^ We used data from UKB under research application #71033.

The Brazilian Longitudinal Study of Adult Health (ELSA-Brasil) study, a large multicenter prospective cohort study conducted in Brazil, enrolled,105 community-dwelling adults aged 35-74 years at their baseline visit during 2008-2010.^6,7^ These participants represent active and retired civil servants from six higher education and research institutions in Brazilian state capitals in three geographical regions of the country: Southeast (Belo Horizonte, Rio de Janeiro, São Paulo and Vitória), South (Porto Alegre) and Northeast (Salvador).^8^ The ELSA-Brasil study aimed to investigate the development and progression of chronic diseases and their determinants in the Brazilian adult population. Baseline data were collected using validated instruments, physical examinations, laboratory assessments, and imaging modalities.^6^ Additionally, all participants underwent protocolized 12-lead ECG and echocardiogram.^6,7^ To ascertain exposure status and to identify changes in baseline, ELSA-Brasil participants present for in-person follow-up visits every three to four years. Moreover, telephone interviews occur annually to obtain information on new diagnoses, hospitalization, and death with adjudicated clinical events based on expert medical record review.^6^

**Study Population**

In YNHHS, to identify patients with prevalent heart failure (HF) at the time of ECG, we identified the first recorded encounter for all patients within the EHR and followed for 1 year. Patients with prevalent HF based on either a diagnosis code for HF or an echocardiogram with left ventricular ejection fraction under 50% or left ventricular diastolic dysfunction (defined as “moderate” or “severe” left ventricular diastolic dysfunction) we excluded from the study. The baseline ECG for patients was defined as an outpatient ECG recorded after this 1-year blanking period to exclude prevalent HF (e**Figure 1**). The YNHHS cohort also excluded patients previously included in the development of the AI-ECG algorithm and a small proportion of individuals who opted out of research participation (<0.01% of all YNHHS patients).

**Study Exposure**

The model development population consisted of 503,516 ECGs from 110,228 unique patients (**Figure 1**). We used raw voltage data from 12-lead ECGs obtained as standard 10-second 12-lead ECGs at a sampling frequency of 500 Hz or 250 Hz and extracted the lead I waveforms. Median filtering was conducted by subtracting a one-second median filter from the acquired signals to eliminate baseline drift. To incorporate noise during the model development, we isolated four distinct noises from a 5-minute random Gaussian noise within four frequency ranges of 3-12 Hz, 12-50 Hz, 50-100 Hz, and 100-150 Hz, each corresponding to the frequency range of a specific type of real-world noise.^9^ The noise with a frequency range of 3-12 Hz reflects the motion artifact noises attributable to tremors, 50-100 Hz accounts for the electrode contact noise, and 12-50 Hz and 100-150 Hz reflect the lower and higher-frequency muscle noises, respectively. Each ECG in the training set was included twice with different random noises signal-to-noise ratios. This augmentation involved a random type of noise and a random signal-to-noise ratio (SNR). For this purpose, we first randomly selected one of the four abovementioned distinct random Gaussian noises. Finally, the selected noise was introduced to the ECG waveform with a random SNR ranging from 0.5 to 1.25, representing a heavy and a light burden of noise in ECGs, respectively.

The employed convolutional neural network (CNN) architecture comprised an input layer with dimensions of (5000, 1, 1), representing a 10-second, 500 Hz, lead I ECG.1 The input layer was followed by seven 2-dimensional convolutional layers, progressively increasing the number of filters from 16 to 64 while incorporating varying kernel sizes (7x1, 5x1, and 3x1) to capture different levels of feature abstraction. A batch normalization layer, a rectified linear unit (ReLU) activation layer, and a 2-dimensional max-pooling layer with different pool sizes (2x1 and 4x1) followed each convolutional layer. Next, the output of the 7th convolutional layer was used as the input for a fully connected network that included two dense layers. Each dense layer was followed by a batch normalization layer, a ReLU activation layer, and a dropout layer with a rate of 0.5. Finally, the model output was a dense layer with a single class and a sigmoid activation to generate the output probability of the label. The loss function was adjusted by calculating model weights using a class re-weighting approach to ensure that the learning is not impacted by the differential prevalence of positive and negative labels.

We defined a positive AI-ECG screen as a model output probability greater than 0.08, representing the probability threshold at which the model achieved a sensitivity of 90% for detecting LVSD during internal validation. We further defined graded thresholds based on AI-ECG probabilities of 0-0.2, 0.2-0.4, 0.4-0.6, 0.6-0.8, and 0.8-1 to evaluate the association of a higher risk score with HF. Notably, while the model was developed for detecting the cross-sectional signature of LVSD using data from the YNHH alone, it was applied across all YNHHS sites and the population-based cohorts without any further development or fine-tuning for prediction of HF risk.

**Study Outcomes and Covariates**

We identified available demographic characteristics across cohorts, including age at the time of ECG, sex, and self-reported race and ethnicity. Comorbidities, including ischemic heart disease, hypertension, and type 2 diabetes mellitus, were defined using relevant EHR diagnosis codes in YNHHS and UKB (**eTable 1**). Obesity was defined as BMI ≥30 kg/m2.

In ELSA-Brasil, covariates were recorded at the baseline study visit.31 Race was self-classified based on Brazil’s National Bureau of Statistics definition and classified as White, Black, “Pardo”, Asian, or Others.31,32In ELSA-Brasil, HF was identified either by in-person interview or the annual telephonic surveillance and investigated by a designated committee that contacted health providers and requested copies of medical records for all hospitalizations. After investigation, the cardiovascular events were adjudicated by an independent review of two cardiologists. A third senior cardiologist defined the event in case of disagreement.^10^ HF was identified from hospitalization records, based on the presence of a clinical diagnosis of HF, with the individual receiving pharmacological therapy for HF, in addition to any of the following: (1) pulmonary congestion on chest X-ray, (2) reduced ejection fraction or systolic dysfunction observed on cardiac imaging, or (3) preserved ejection fraction with evidence of moderate to severe diastolic dysfunction.

Information about all-cause death was available in the YNHHS EHR, with in-hospital mortality data supplemented from the Connecticut death index to improve capturing out-of-hospital patient mortality. Similarly, information about mortality was available in UKB via linkage to the EHR and the UK national death registries. Information about death in the ELSA-Brasil study was recorded via telephonic surveillance and confirmed using the national mortality database and death certificates.

**Study Comparator**

We employed two established clinical models to predict HF risk, the pooled cohort equations to prevent HF (PCP-HF) and the predicting risk of cardiovascular disease events (PREVENT) equations, as the baseline models.^11–13^ The PCP-HF entails sex- and race-specific equations for estimating the 10-year risk of incident HF. However, the PREVENT predicts the 10-year HF risk independently of race. To align with the score development across cohorts, the PCP-HF score was calculated for White and Black individuals between 30 and 80 years of age, and the PREVENT score was computed for all individuals between 30 and 80 years of age with complete documentation of the score covariates. The calculated 10-year risk score was adjusted based on the length of follow-up for each individual to estimate the risk of HF over the study period.

In YNHHS, PCP-HF and PREVENT features were extracted from the EHR. Body mass index (BMI), systolic blood pressure, and laboratory measurements closest to and within two years of the ECG acquisition date were used for calculation. In UKB, the demographic features were identified from the baseline visit. Blood pressure measurement and smoking status assessment were conducted at the time of ECG acquisition. Laboratory values were measured in the first and second study visits, while ECGs were recorded in third and fourth visits.^14^ We used the laboratory values closest to the ECG acquisition for the calculation of the PCP-HF and PREVENT scores. History of hypertension and diabetes were defined using ICD diagnosis codes from the linked EHR and self-reported use of anti-hypertensive and anti-hyperglycemic medications was recorded at the time of ECG acquisition.^4^ In ELSA-Brasil, all features, including the ECG recording, were captured at the baseline visit using established study protocols.^15,16^

**Statistical Analysis**

Integrated discrimination improvement (IDI) was calculated as the difference between the improvements in the average predicted probabilities for those with and without the outcome for the AI-ECG model output vs. PCP-HF and PREVENT scores in each data source. Categorical net reclassification improvement (NRI) was calculated for the 0.08 threshold of the AI-ECG model. We also calculated event and non-event NRIs. Sensitivity, specificity, positive predictive value (PPV), and negative predictive value (NPV) of AI-ECG, PCP-HF, and PREVENT for predicting new-onset HF were also reported with censoring the observations at the median duration of follow-up. Net benefit evaluates true positives while accounting for the potential for increased false positives, ranging from 0-1, with higher values showing greater benefit. This was calculated using the following formula:

$$Net Benefit = \left( \frac{True Positive Rate-False Positive Rate}{1-Probability Threshold} \right)$$

Categorical variables were reported as counts (percentages), and continuous variables as median (interquartile range [IQR]). All statistical tests were 2-sided with a level of significance set at 0.05. Harrell’s C-statistic was calculated based on various Cox proportional hazard models with the AI-ECG model probability, age, sex, PCP-HF, and PREVENT scores as covariates. In Cox proportional hazard models, we treated death as a censoring event or, in sensitivity analyses, included death as part of a composite outcome in the dependent variable. compareC package in R was used for calculating and comparing Harrel's C-statistics.^17^ We also evaluated the association of AI-ECG with HF, overall and across racial groups, after adjusting for the PCP-HF score in Cox models. All analyses were conducted using a combination of Python 3.11.2 and R version 4.2.0. The Yale Institutional Review Board approved the study protocol and waived the need for informed consent as the study involves secondary analysis of pre-existing data.

**eFigure 1. Overview of Cohort Creation at the Yale New Haven Health System.** Abbreviations: ECG, Electrocardiograms; EF, Ejection Fraction; HF, Heart Failure; YNHHS, Yale New Haven Health System.

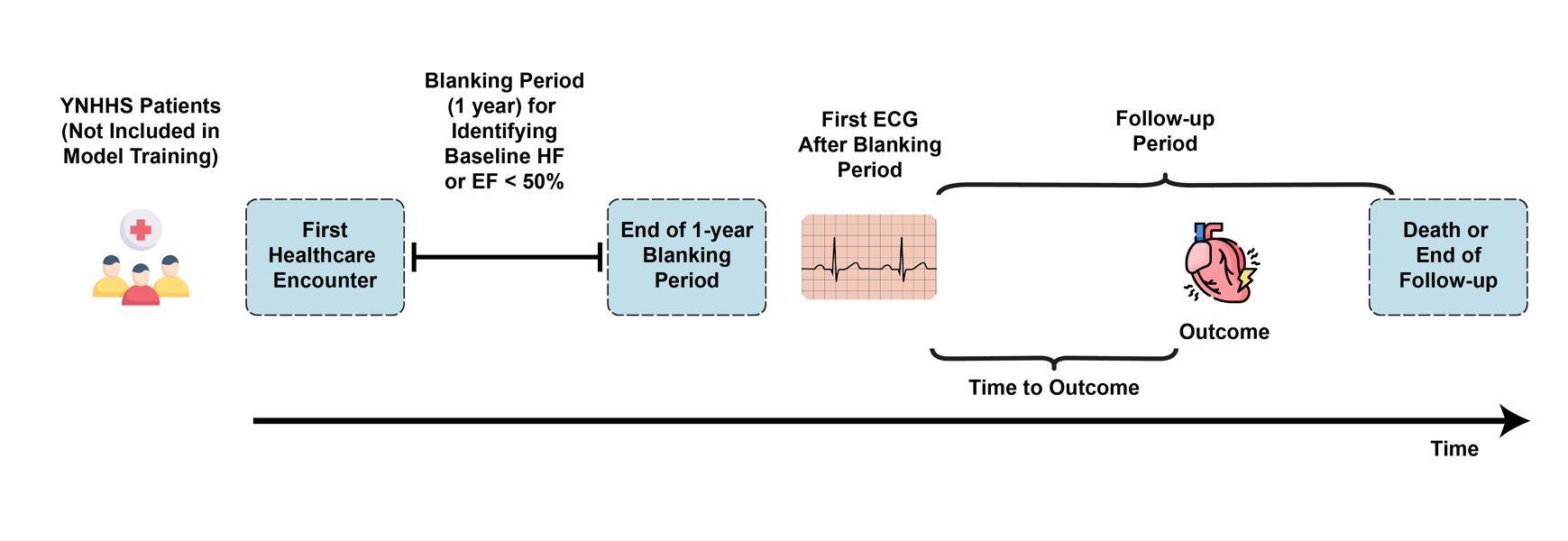

**eFigure 2. Consort Diagram for Study Cohorts.** Abbreviations: ECG, Electrocardiogram; ELSA Brasil, Brazilian Longitudinal Study of Adult Health; HF, Heart Failure; YNHHS, Yale New Haven Health System

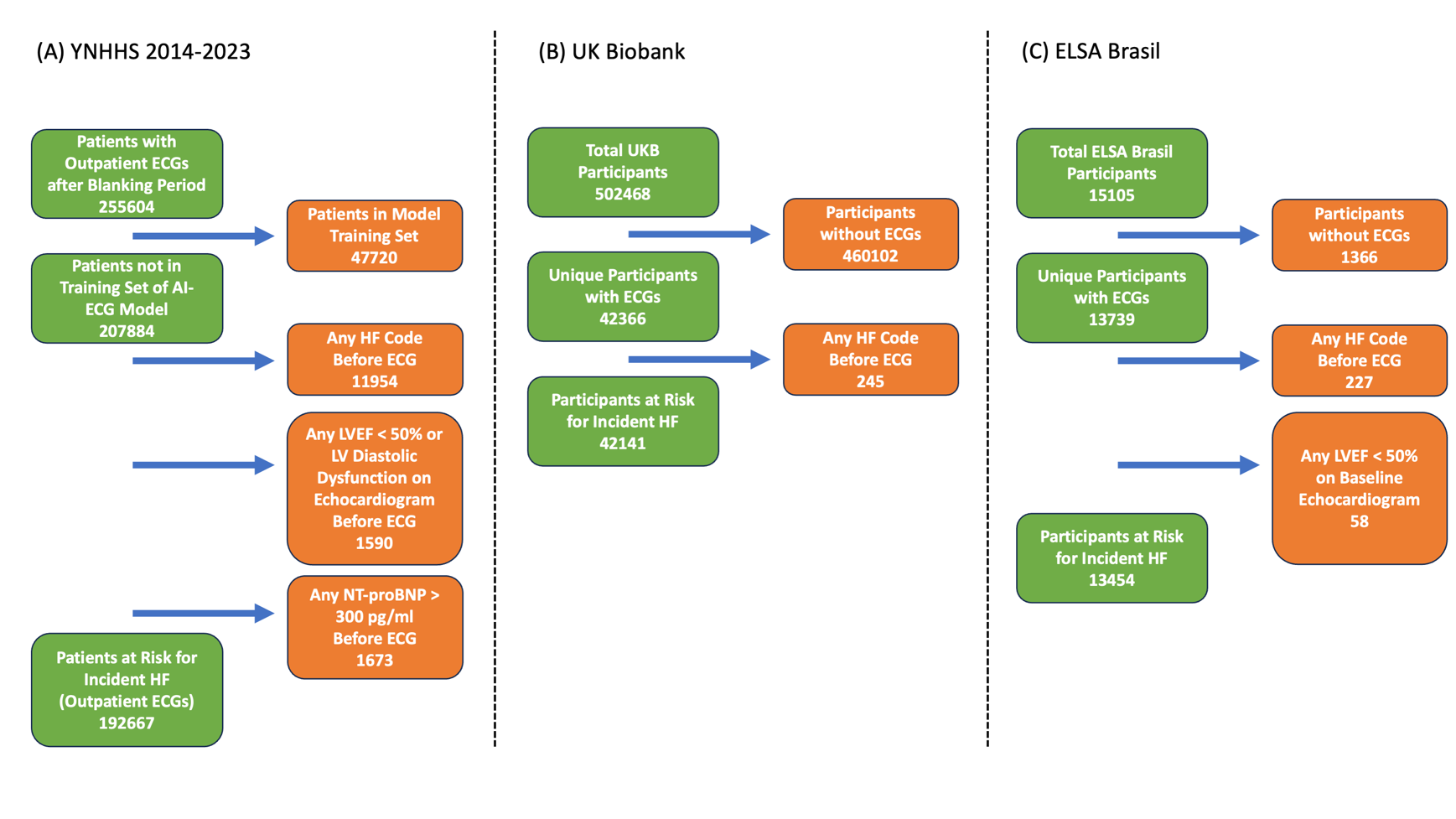

**eFigure 3. Model Performance for Cross-sectional Detection of Left Ventricular Systolic Dysfunction in the Yale New Haven Hospital Held-out Test Set and Across External Validation Cohorts.** Abbreviations: AUROC, area under the receiver operating characteristic curve; CI, confidence interval; YNHH, Yale New Haven Hospital.

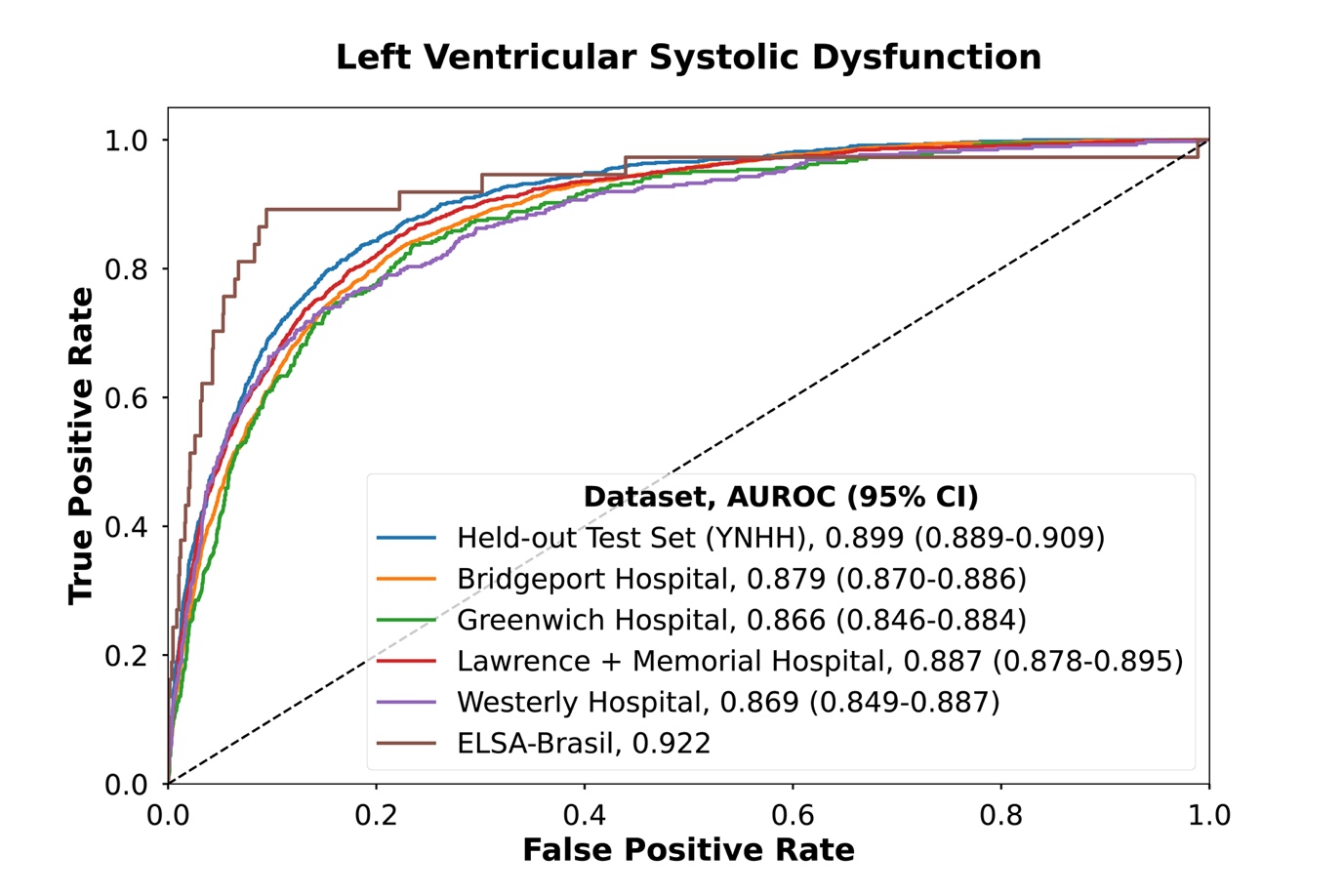

**eFigure 4. Age- and Sex-adjusted Hazard for Heart Failure across Model Output Probability Bins.** Abbreviations: ELSA-Brasil, Brazilian Longitudinal Study of Adult Health; UKB, UK Biobank; YNHHS, Yale New Haven Health System.

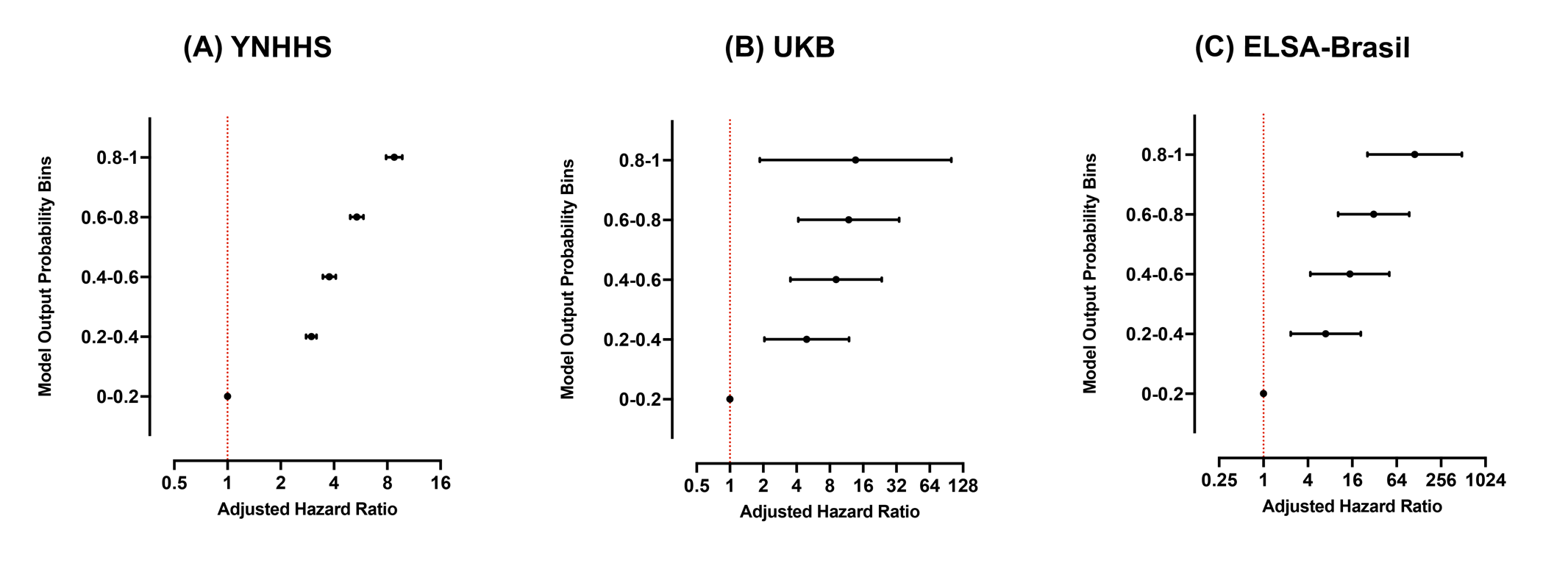

**eFigure 5. Net Benefit of AI-ECG Model Output Probability and Pooled Cohort Equations to Prevent Heart Failure) and Predicting Risk of Cardiovascular Disease Events Equations for Predicting Incident Heart Failure Across Thresholds at (A) Yale New Haven Health System (B) UK Biobank (C) Brazilian Longitudinal Study of Adult Health.** Abbreviations: AI-ECG, Artificial Intelligence-enhanced Electrocardiography; ELSA-Brasil, Brazilian Longitudinal Study of Adult Health; PCP-HF, Pooled Cohort Equations to Prevent Heart Failure; PREVENT, Predicting Risk of Cardiovascular Disease Events; UKB, UK Biobank; YNHHS, Yale New Haven Health System

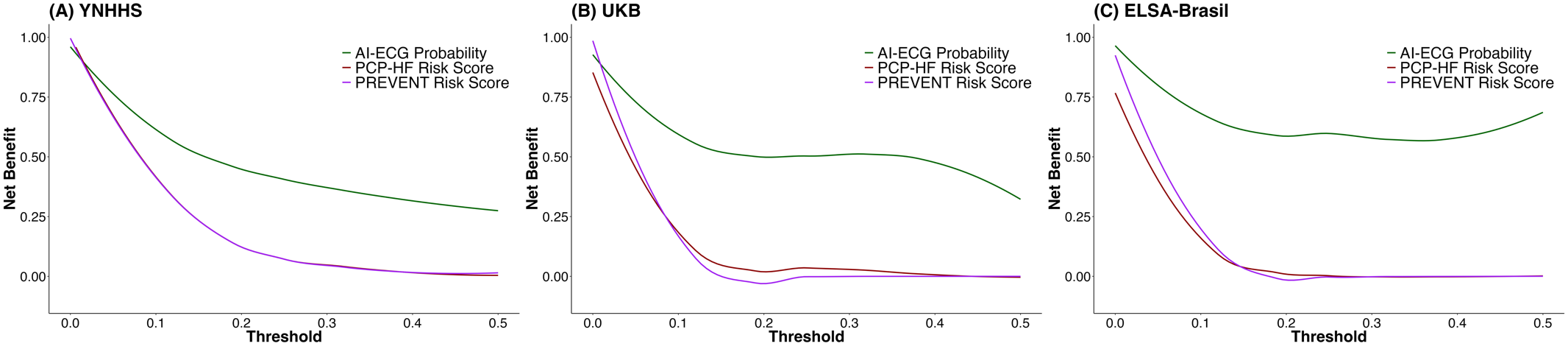

**eFigure 6. Cumulative Hazard for Heart Failure Adjusted for Pooled Cohort Equations to Prevent Heart Failure (PCP-HF) Risk Score at (A) Yale New Haven Health System (B) UK Biobank (C) Brazilian Longitudinal Study of Adult Health.** Abbreviations: aHR, Adjusted Hazard Ratio; ELSA-Brasil, Brazilian Longitudinal Study of Adult Health; UKB, UK Biobank; YNHHS, Yale New Haven Health System.

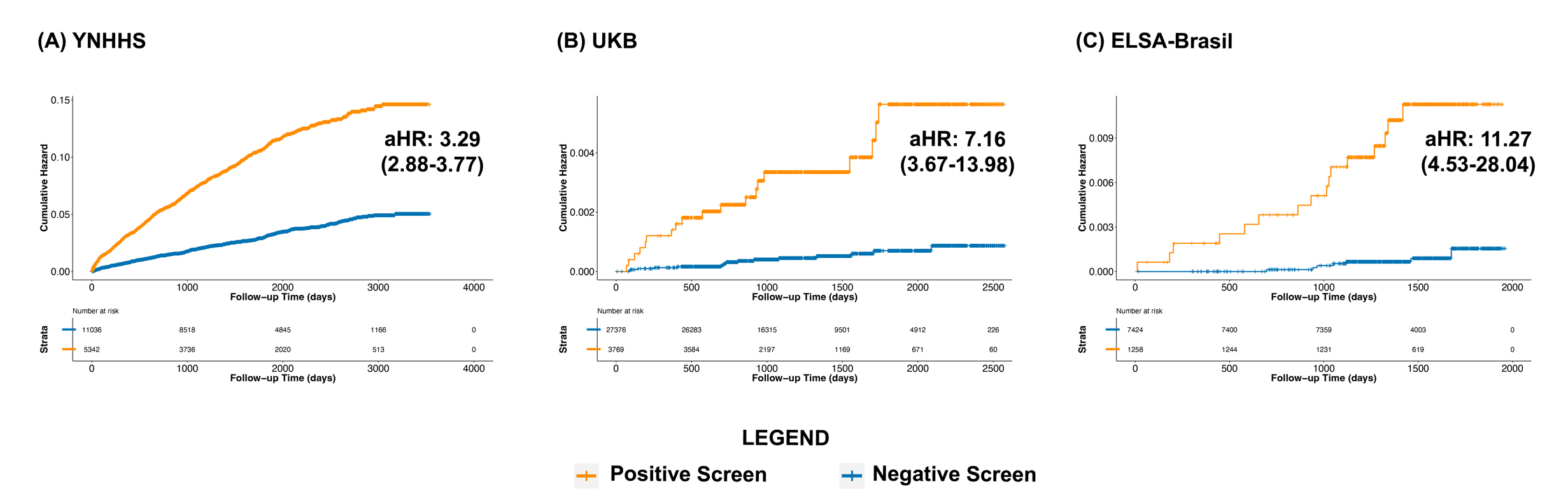

**eFigure 7. Cumulative Hazard for Heart Failure Adjusted for Pooled Cohort Equations to Prevent Heart Failure (PCP-HF) Risk Score at Yale New Haven Health System.** Abbreviations: aHR, Adjusted Hazard Ratio

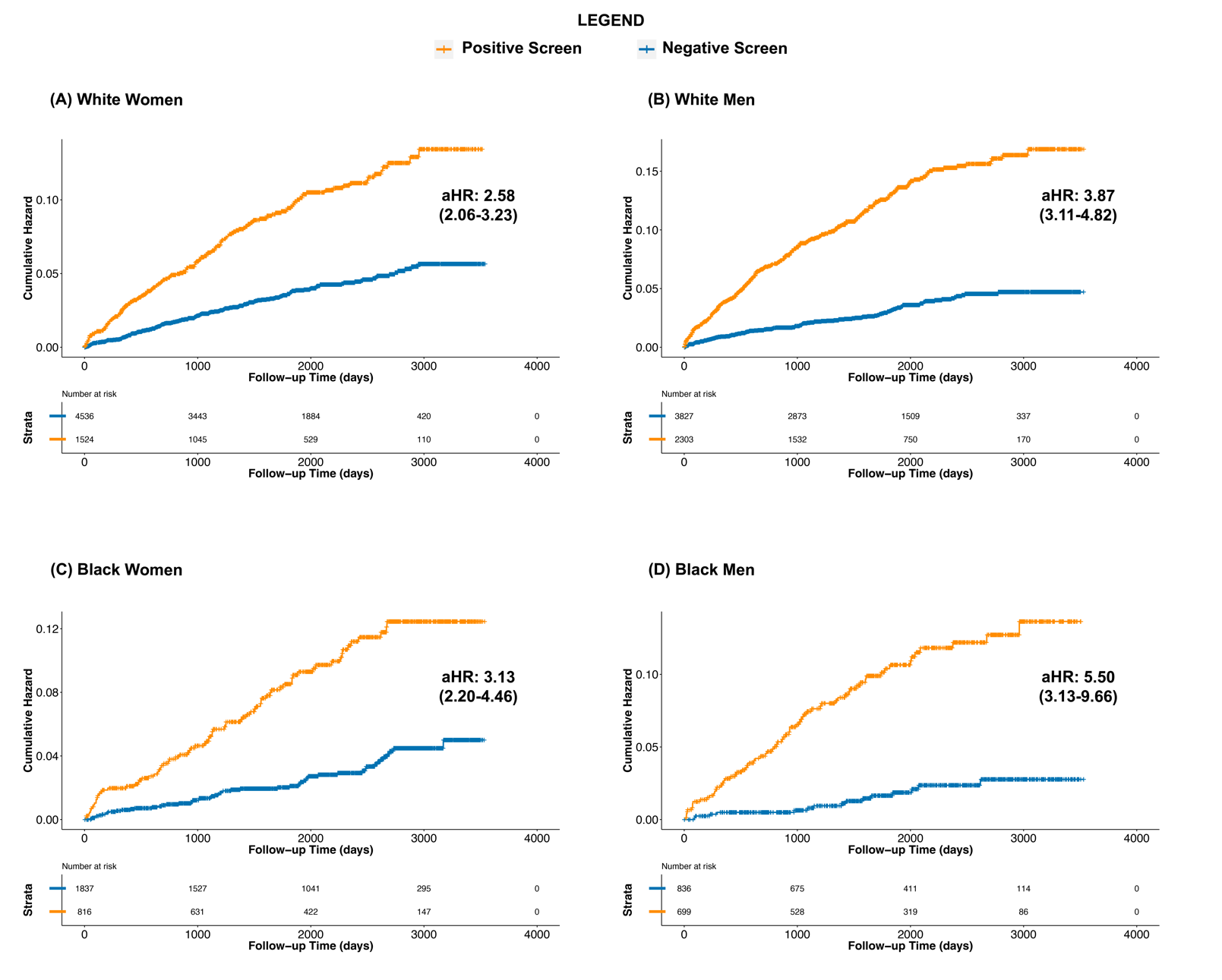

**eTable 1. International Classification of Disease Tenth Revision Codes for the Identification of Comorbidities and Outcomes.** Abbreviations: ICD-10-CM, International Classification of Disease Tenth Revision Clinical Modification Codes.

| **Condition** | **ICD-10-CM codes** |
| --- | --- |
| **Heart Failure** | 'I11.0','I13.0','I13.2','I50','I50.0','I50.1','I50.9','Z95.81','I09.81' |
| **Acute Myocardial Infarction** | 'I21', 'I22', 'I23', 'I24.0', 'I24.8', 'I24.9' |
| **Stroke** | 'G45','G45.0','G45.1','G45.2','G45.3','G45.4','G45.8','G45.9',  'I63','I63.0','I63.1','I63.2','I63.3','I63.4','I63.5','I63.8','I63.9','I64',  'I65','I65.0','I65.1','I65.2','I65.3','I65.8','I65.9','I66','I66.0','I66.1',  'I66.2','I66.3','I66.4','I66.8','I66.9','I67.2','I69.3','I69.4' |
| **Type 2 Diabetes Mellitus** | 'E11','E11.0','E11.1','E11.2','E11.3','E11.4','E11.5','E11.6',  'E11.7','E11.8','E11.9','O24.1' |
| **Hypertension** | 'I10','I11','I11.0','I11.9','I12','I12.0','I12.9',  'I13','I13.0','I13.1','I13.2','I13.9','I67.4',  'O10','O10.0','O10.1','O10.2','O10.3','O10.9','O11' |
| **Ischemic Heart Disease** | 'I20', 'I20.0', 'I20.8', 'I20.9', 'I21', 'I21.0', 'I21.1', 'I21.2', 'I21.3',  'I21.4', 'I21.9', 'I21.X', 'I22', 'I22.0', 'I22.1', 'I22.8', 'I22.9', 'I23', 'I23.0', 'I23.1', 'I23.2', 'I23.3', 'I23.4', 'I23.5', 'I23.6', 'I23.8', 'I24', 'I24.0', 'I24.1', 'I24.8', 'I24.9', 'I25', 'I25.0', 'I25.1', 'I25.2', 'I25.5', 'I25.6', 'I25.8', 'I25.9', 'Z95.1', 'Z95.5' |

**eTable 2. Model Performance Measures for Cross-sectional Detection of Left Ventricular Systolic Dysfunction in the Yale New Haven Hospital Held-out Test Set and Across External Validation Cohorts.** Abbreviations: AUROC, area under the receiver operating characteristic curve; NPV, negative predictive value; OR, odds ratio; PPV, positive predictive value.

| **Dataset** | **Total Number** | **Diagnostic OR** | **AUROC** | **F1 Score** | **Prevalence** | **Sensitivity** | **Specificity** | **PPV** | **NPV** |
| --- | --- | --- | --- | --- | --- | --- | --- | --- | --- |
| **Held-out Test Set** | | | | | | | | | |
| **Yale New Haven Hospital** | 10860 | 24.9 (19.7-31.5) | 0.899 (0.889-0.909) | 0.346 | 7.6% | 90.3% (89.7-90.8) | 72.9% (72.1-73.8) | 21.4% (20.6-22.2) | 98.9% (98.7-99.1) |
| **External Validation Sites** | | | | | | | | | |
| **Bridgeport Hospital** | 17915 | 19.2 (16.2-22.8) | 0.879 (0.870-0.886) | 0.353 | 9.9% | 91.5% (91.1-91.9) | 64.2% (63.4-64.9) | 21.9% (21.3-22.5) | 98.6% (98.4-98.7) |
| **Greenwich Hospital** | 4306 | 15.6 (11.3-21.4) | 0.866 (0.846-0.884) | 0.334 | 8.5% | 87.8% (86.8-88.8) | 68.4% (67.0-69.8) | 20.6% (19.4-21.8) | 98.4% (98.0-98.7) |
| **Lawrence + Memorial Hospital** | 17730 | 21.4 (17.9-25.6) | 0.887 (0.878-0.895) | 0.338 | 8.3% | 90.7% (90.2-91.1) | 68.8% (68.1-69.5) | 20.8% (20.2-21.4) | 98.8% (98.6-99.0) |
| **Westerly Hospital** | 3614 | 13.9 (10.2-19.0) | 0.869 (0.849-0.887) | 0.371 | 10.7% | 87.8% (86.8-88.9) | 65.8% (64.3-67.4) | 23.5% (22.1-24.9) | 97.8% (97.4-98.3) |
| **ELSA-Brasil** | 3012 | 51.1 (22.2-117.7) | 0.922 | 0.201 | 1.2% | 81.1% (79.7-82.5) | 92.3% (91.3-93.2) | 11.5% (10.4-12.7) | 99.7% (99.6-99.9) |

**eTable 3. Population Characteristics of the Yale New Haven Health System Sites.** Abbreviations: AMI, acute myocardial infarction; ECG, Electrocardiogram; HF, heart failure; IQR, Interquartile Range; LVEF, Left Ventricular Ejection Fraction; NEMG, Northeast Medical Group; L&M, Lawrence and Memorial Hospital; YNHH, Yale New Haven Hospital

| **Characteristic** | | **YNHH** | **Bridgeport** | **Greenwich** | **L&M** | **Westerly** | **NEMG** |
| --- | --- | --- | --- | --- | --- | --- | --- |
| **Number** | | 96317 | 32377 | 17746 | 19080 | 4529 | 22618 |
| **Age at ECG, Median [IQR]** | | 54.3 [38.6,67.1] | 51.4 [37.3,64.0] | 56.0 [42.6,70.2] | 58.3 [43.5,71.0] | 65.5 [53.8,76.0] | 64.1 [54.3,73.7] |
| **Female Sex, N (%)** | | 55580 (57.7) | 19669 (60.7) | 10406 (58.6) | 11235 (58.9) | 2464 (54.4) | 11827 (52.3) |
| **Race/Ethnicity,**  **N (%)** | **White** | 57231 (59.4) | 12469 (38.5) | 12081 (68.1) | 13579 (71.2) | 4185 (92.4) | 18312 (81.0) |
|  | **Black** | 18293 (19.0) | 8232 (25.4) | 969 (5.5) | 1726 (9.0) | 81 (1.8) | 1322 (5.8) |
|  | **Hispanic** | 14942 (15.5) | 10291 (31.8) | 3580 (20.2) | 2723 (14.3) | 121 (2.7) | 1599 (7.1) |
|  | **Asian** | 2033 (2.1) | 348 (1.1) | 509 (2.9) | 266 (1.4) | 34 (0.8) | 363 (1.6) |
|  | **Other** | 1138 (1.2) | 309 (1.0) | 137 (0.8) | 376 (2.0) | 57 (1.3) | 142 (0.6) |
|  | **Missing** | 2680 (2.8) | 728 (2.2) | 470 (2.6) | 410 (2.1) | 51 (1.1) | 880 (3.9) |
| **Death, N (%)** | | 5082 (5.3) | 1587 (4.9) | 811 (4.6) | 1161 (6.1) | 381 (8.4) | 1359 (6.0) |
| **Follow-up Time, Years; Median [IQR]** | | 4.9 [2.7,6.9] | 4.8 [3.2,6.6] | 5.3 [3.6,6.8] | 3.4 [2.0,4.7] | 3.3 [1.5,4.6] | 4.7 [3.0,6.7] |
| **Positive Screens, N (%)** | | 20670 (21.5) | 6930 (21.4) | 3227 (18.2) | 4597 (24.1) | 1354 (29.9) | 5997 (26.5) |
| **Hypertension at baseline, N (%)** | | 42576 (44.2) | 13411 (41.4) | 6235 (35.1) | 9390 (49.2) | 2722 (60.1) | 13881 (61.4) |
| **Type-2 diabetes mellitus at baseline, N (%)** | | 16901 (17.5) | 6356 (19.6) | 2345 (13.2) | 3846 (20.2) | 1402 (31.0) | 4672 (20.7) |
| **Obesity at baseline, N (%)** | | 16543 (17.2) | 4950 (15.3) | 1262 (7.1) | 3316 (17.4) | 1178 (26.0) | 3244 (14.3) |
| **Atrial fibrillation at baseline, N (%)** | | 2036 (2.1) | 479 (1.5) | 407 (2.3) | 621 (3.3) | 229 (5.1) | 974 (4.3) |
| **Left bundle branch block at baseline, N (%)** | | 1212 (1.3) | 227 (0.7) | 151 (0.9) | 326 (1.7) | 98 (2.2) | 383 (1.7) |
| **Use of antihypertensive drugs at baseline, N (%)** | | 22806 (23.7) | 7549 (23.3) | 2560 (14.4) | 4815 (25.2) | 1828 (40.4) | 8053 (35.6) |
| **Use of antihyperglycemic drugs at baseline, N (%)** | | 14584 (15.1) | 5650 (17.5) | 1652 (9.3) | 3355 (17.6) | 1089 (24.0) | 4190 (18.5) |
| **End-stage renal disease, N (%)** | | 329 (0.3) | 79 (0.2) | 24 (0.1) | 60 (0.3) | 15 (0.3) | 40 (0.2) |
| **Primary HF hospitalization during follow-up, N (%)** | | 1454 (1.5) | 594 (1.8) | 305 (1.7) | 526 (2.8) | 231 (5.1) | 587 (2.6) |
| **Primary HF hospitalization or an echocardiogram with LVEF < 50% during follow-up, N (%)** | | 2904 (3.0) | 1185 (3.7) | 460 (2.6) | 990 (5.2) | 336 (7.4) | 1639 (7.2) |
| **Any HF hospitalization during follow-up, N (%)** | | 6313 (6.6) | 2079 (6.4) | 917 (5.2) | 1443 (7.6) | 602 (13.3) | 2351 (10.4) |
| **Any HF hospitalization or an echocardiogram with LVEF < 50% during follow-up, N (%)** | | 7047 (7.3) | 2374 (7.3) | 995 (5.6) | 1689 (8.9) | 656 (14.5) | 2944 (13.0) |
| **Primary AMI hospitalization during follow-up, N (%)** | | 50 (0.1) | 47 (0.1) | 16 (0.1) | 37 (0.2) | 22 (0.5) | 194 (0.9) |
| **Primary Stroke hospitalization during follow-up, N (%)** | | 1210 (1.3) | 539 (1.7) | 370 (2.1) | 529 (2.8) | 160 (3.5) | 473 (2.1) |
| **Major Adverse Cardiovascular Events during follow-up, N (%)** | | 7162 (7.4) | 2513 (7.8) | 1367 (7.7) | 1999 (10.5) | 682 (15.1) | 2316 (10.2) |

**eTable 4. Model Performance for Predicting Heart Failure Risk.** Abbreviations: IHD, Ischemic Heart Disease; HTN, hypertension; NEMG, Northeast Medical Group; L&M, Lawrence and Memorial Hospital; T2DM, type-2 diabetes mellitus; YNHH, Yale New Haven Hospital

| **Model** | **Covariates** | **YNHH** | | **Bridgeport** | | **Greenwich** | | **L&M** | | **Westerly** | | **NEMG** | |
| --- | --- | --- | --- | --- | --- | --- | --- | --- | --- | --- | --- | --- | --- |
|  |  | **Positive Screen** | **Per 0.1 Increment** | **Positive Screen** | **Per 0.1 Increment** | **Positive Screen** | **Per 0.1 Increment** | **Positive Screen** | **Per 0.1 Increment** | **Positive Screen** | **Per 0.1 Increment** | **Positive Screen** | **Per 0.1 Increment** |
| **Cox Proportional Hazard Model** | Model Probability | 4.98 (4.49-5.52) | 1.47 (1.44-1.50) | 5.17 (4.39-6.08) | 1.46 (1.42-1.50) | 5.52 (4.41-6.92) | 1.54 (1.48-1.61) | 4.43 (3.72-5.27) | 1.43 (1.39-1.48) | 4.17 (3.19-5.44) | 1.34 (1.28-1.41) | 4.45 (3.76-5.25) | 1.37 (1.33-1.41) |
| **Cox Proportional Hazard Model** | Model Probability + Age + Sex | 3.47 (3.12-3.87) | 1.34 (1.31-1.37) | 3.30 (2.79-3.91) | 1.33 (1.28-1.37) | 2.82 (2.23-3.57) | 1.33 (1.26-1.39) | 2.92 (2.44-3.49) | 1.31 (1.27-1.36) | 3.09 (2.36-4.05) | 1.24 (1.18-1.31) | 3.08 (2.59-3.65) | 1.27 (1.23-1.31) |
| **Cox Proportional Hazard Model** | Model Probability + Age + Sex + IHD + HTN + T2DM + Obesity | 2.97 (2.67-3.31) | 1.30 (1.28-1.33) | 2.73 (2.31-3.24) | 1.28 (1.24-1.32) | 2.46 (1.95-3.10) | 1.27 (1.21-1.34) | 2.46 (2.05-2.94) | 1.28 (1.24-1.33) | 2.80 (2.13-3.68) | 1.21 (1.15-1.28) | 2.69 (2.27-3.20) | 1.25 (1.21-1.29) |
| **Fine-Gray Subdistribution Hazard Model** | Model Probability + Age + Sex + Competing Risk of Death | 3.38 (3.09-3.69) | 1.32 (1.30-1.34) | 3.30 (2.92-3.73) | 1.31 (1.28-1.34) | 2.88 (2.45-3.37) | 1.28 (1.25-1.32) | 3.01 (2.63-3.45) | 1.30 (1.27-1.33) | 3.04 (2.47-3.74) | 1.23 (1.18-1.27) | 2.96 (2.49-3.52) | 1.26 (1.22-1.30) |
| **Fine-Gray Subdistribution Hazard Model** | Model Probability Age + Sex + IHD + HTN + T2DM + Obesity + Competing Risk of Death | 2.93 (2.62-3.27) | 1.30 (1.27-1.32) | 2.71 (2.28-3.21) | 1.27 (1.23-1.31) | 2.39 (1.90-3.00) | 1.24 (1.18-1.30) | 2.32 (1.94-2.78) | 1.26 (1.22-1.31) | 2.86 (2.19-3.72) | 1.20 (1.14-1.27) | 2.57 (2.16-3.06) | 1.23 (1.19-1.27) |

**eTable 5. Model Performance for Prediction of Heart Failure Across Demographic Subgroups.** Abbreviations: CI, Confidence Interval; ELSA-Brasil, Brazilian Longitudinal Study of Adult Health; HF, Heart Failure; UKB, UK Biobank; YNHHS, Yale New Haven Health System

| **Subgroup** | | **YNHHS** | | | **UKB** | | | **ELSA-Brasil** | | |
| --- | --- | --- | --- | --- | --- | --- | --- | --- | --- | --- |
|  |  | **Total Number of Individuals at Risk** | **Number of HF Events** | **Age- and Sex- Adjusted Cox Proportional Hazard Ratios (95% CI)** | **Total Number of Individuals at Risk** | **Number of HF Events** | **Age- and Sex- Adjusted Cox Proportional Hazard Ratios (95% CI)** | **Total Number of Individuals at Risk** | **Number of HF Events** | **Age- and Sex- Adjusted Cox Proportional Hazard Ratios (95% CI)** |
| **Age < 65** | | 131335 (68.2) | 1061 | 4.84 (4.28-5.47) | 20802 (49.4) | 9 | 6.30 (1.68-23.57) | 12038 (89.4) | 21 | 11.41 (4.55-28.64) |
| **Age ≥ 65** | | 61332 (31.8) | 2636 | 2.80 (2.59-3.03) | 21345 (50.6) | 37 | 5.86 (3.06-11.23) | 1416 (10.6) | 10 | 4.69 (1.31-16.71) |
| **Female** | | 111181 (57.7) | 1885 | 2.95 (2.69-3.24) | 21795 (51.7) | 11 | 3.76 (1.10-12.88) | 7348 (54.6) | 11 | 10.21 (3.88-26.91) |
| **Male** | | 81486 (42.3) | 1812 | 3.75 (3.40-4.15) | 20346 (48.3) | 35 | 6.92 (3.52-13.60) | 6106 (45.4) | 20 | 6.76 (2.03-22.47) |
| **Race/ Ethnicity** | **White** | 117857 (61.2) | 2769 | 3.02 (2.80-3.27) | 40691 (96.6) | 46 | 5.96 (3.32-10.68) | 6920 (51.4) | 15 | 10.17 (3.40-30.46) |
|  | **Black** | 30623 (15.9) | 470 | 4.36 (3.48-5.44) | 304 (0.7) | 0 | - | 2130 (15.8) | 9 | 11.80 (2.38-58.47) |
|  | **Hispanic** | 33256 (17.3) | 338 | 4.40 (3.54-5.47) | 0 | - | - | - | - | - |
|  | **Asian** | 3553 (1.8) | 32 | 5.21 (2.50-10.87) | 600 (1.4) | 0 | - | - | - | - |
|  | **Other** | 2159 (1.1) | 28 | 2.98 (1.39-6.39) | 546 (1.3) | 0 | - | 637 (4.7) | 0 | - |
|  | **Brazilian “Pardo”** | - | - | - | - | - | - | 3767 (28.0) | 7 | 3.69 (0.81-16.8) |
|  | **Missing** | 5219 (2.7) | 60 | 2.34 (1.38-3.97) | 0 | - | - | - | - | - |

**eTable 6. Age- and Sex- Adjusted Cox Proportional Hazard Models for the Prediction of Heart Failure-related Outcomes.** Abbreviations: AMI, acute myocardial infarction; ELSA Brasil, Brazilian Longitudinal Study of Adult Health; HF, heart failure; LVEF, Left Ventricular Ejection Fraction; UKB, UK Biobank; YNHHS, Yale New Haven Health System

| **Outcome** | **YNHHS** | **UKB** | **ELSA-Brasil** |
| --- | --- | --- | --- |
| **Primary HF Hospitalization** | 3.31 (3.10-3.54) | 5.96 (3.32-10.68) | 8.74 (4.13-18.48) |
| **Primary HF Hospitalization or an Echocardiogram with LVEF < 50%** | 3.87 (3.69-4.06) | - | - |
| **Primary HF Hospitalization or Death** | 1.97 (1.90-2.04) | 1.91 (1.51-2.41) | 2.59 (1.99-3.35) |
| **Any Hospitalization with HF** | 2.43 (2.34-2.51) | 3.55 (2.72-4.64) | - |
| **Any Hospitalization with HF or an Echocardiogram with LVEF < 50%** | 2.62 (2.54-2.70) | - | - |
| **Any HF Hospitalization or Death** | 2.06 (2.01-2.12) | 2.18 (1.81-2.63) | - |

**eTable 7. Age- and Sex- Adjusted Cox Proportional Hazard Models for the Prediction of Heart Failure-related Outcomes.** Abbreviations: AMI, acute myocardial infarction; HF, heart failure; NEMG, Northeast Medical Group; LVEF, Left Ventricular Ejection Fraction; L&M, Lawrence and Memorial Hospital; YNHH, Yale New Haven Hospital

| **Outcome** | **YNHH** | **Bridgeport** | **Greenwich** | **L&M** | **Westerly** | **NEMG** |
| --- | --- | --- | --- | --- | --- | --- |
| **Primary HF Hospitalization** | 4.98 (4.49-5.52) | 3.30 (2.79-3.91) | 2.82 (2.23-3.57) | 2.92 (2.44-3.49) | 3.09 (2.36-4.05) | 3.08 (2.59-3.65) |
| **Primary HF Hospitalization or an Echocardiogram with LVEF < 50%** | 3.76 (3.49-4.06) | 3.61 (3.20-4.07) | 2.76 (2.28-3.34) | 3.46 (3.04-3.95) | 2.71 (2.17-3.37) | 4.86 (4.37-5.41) |
| **Primary HF Hospitalization or Death** | 1.89 (1.79-1.99) | 1.81 (1.65-1.98) | 1.97 (1.74-2.23) | 2.20 (1.99-2.44) | 1.75 (1.48-2.08) | 2.09 (1.9-2.30) |
| **Any Hospitalization with HF** | 2.39 (2.28-2.52) | 2.47 (2.26-2.70) | 2.17 (1.89-2.48) | 2.69 (2.42-2.99) | 2.00 (1.70-2.35) | 2.25 (2.07-2.45) |
| **Any Hospitalization with HF or an Echocardiogram with LVEF < 50%** | 2.51 (2.39-2.63) | 2.56 (2.36-2.78) | 2.19 (1.93-2.50) | 2.83 (2.57-3.13) | 1.97 (1.68-2.30) | 2.87 (2.67-3.09) |
| **Any HF Hospitalization or Death** | 2.01 (1.93-2.10) | 1.99 (1.85-2.14) | 1.96 (1.76-2.18) | 2.31 (2.12-2.51) | 1.74 (1.52-2.00) | 2.05 (1.91-2.21) |

**eTable 8. Age- and Sex-Adjusted Cox Proportional Hazard Models for the Prediction of Heart Failure across Model Output Probabilities.** Abbreviations: ELSA Brasil, Brazilian Longitudinal Study of Adult Health; UKB, UK Biobank; YNHHS, Yale New Haven Health System

| **Model output probability bins** | **YNHHS** | **UKB** | **ELSA-Brasil** |
| --- | --- | --- | --- |
| **0-0.2** | Reference | Reference | Reference |
| **0.2-0.4** | 2.91 (2.65-3.19) | 4.94 (2.05-11.93) | 6.98 (2.34-20.82) |
| **0.4-0.6** | 3.55 (3.15-3.99) | 9.11 (3.52-23.57) | 14.83 (4.32-50.95) |
| **0.6-0.8** | 4.88 (4.29-5.56) | 11.87 (4.15-33.9) | 31.17 (10.27-94.66) |
| **0.8-1.0** | 7.49 (6.33-8.87) | 13.68 (1.86-100.45) | 112.33 (25.75-489.96) |

**eTable 9. Age- and Sex-Adjusted Cox Proportional Hazard Models for the Prediction of Heart Failure across Model Output Probabilities at the Yale New Haven Health System Sites.** Abbreviations: NEMG, Northeast Medical Group; L&M, Lawrence and Memorial Hospital; YNHH, Yale New Haven Hospital

| **Model output probability bins** | **YNHH** | **Bridgeport** | **Greenwich** | **L&M** | **Westerly** | **NEMG** |
| --- | --- | --- | --- | --- | --- | --- |
| **0-0.2** | Reference | Reference | Reference | Reference | Reference | Reference |
| **0.2-0.4** | 3.58 (3.10-4.14) | 2.20 (1.72-2.83) | 2.63 (1.90-3.64) | 2.34 (1.81-3.02) | 3.12 (2.25-4.33) | 2.34 (1.86-2.94) |
| **0.4-0.6** | 3.67 (3.03-4.45) | 3.97 (2.95-5.35) | 3.85 (2.55-5.82) | 3.55 (2.65-4.76) | 2.12 (1.22-3.69) | 3.08 (2.30-4.13) |
| **0.6-0.8** | 5.65 (4.63-6.91) | 5.08 (3.60-7.15) | 4.56 (2.80-7.43) | 4.34 (3.10-6.07) | 3.80 (2.30-6.26) | 3.79 (2.73-5.24) |
| **0.8-1.0** | 9.14 (7.05-11.85) | 6.44 (4.17-9.94) | 8.35 (4.10-17.0) | 9.59 (6.40-14.36) | 3.12 (1.27-7.63) | 5.15 (3.36-7.90) |

**eTable 10. Categorical and Continuous Time-to-Event, Event and Non-event Net Reclassification Index of AI-ECG Model Output Probability over Pooled Cohort Equations to Prevent Heart Failure, and Predicting Risk of Cardiovascular Disease Events Equations.** Abbreviations: ELSA-Brasil, Brazilian Longitudinal Study of Adult Health; NRI, Net Reclassification Index; PCP-HF, Pooled Cohort equations to Prevent Heart Failure; PREVENT, Predicting Risk of Cardiovascular Disease Events; UKB, UK Biobank; YNHHS, Yale New Haven Health System.

| **Metric** | **YNHHS** | | **UKB** | | **ELSA-Brasil** | |
| --- | --- | --- | --- | --- | --- | --- |
| **Comparator** | **PCP-HF** | **PREVENT** | **PCP-HF** | **PREVENT** | **PCP-HF** | **PREVENT** |
| **Categorical NRI** |  |  |  |  |  |  |
| **NRI** | 0.158 (0.102 to 0.218) | 0.128 (0.092 to 0.167) | 0.291 (0.092 to 0.494) | 0.330 (0.088 to 0.547) | 0.488 (0.188 to 0.741) | 0.363 (0.118 to 0.592) |
| **NRI+** | 0.228 (0.175 to 0.285) | 0.197 (0.162 to 0.236) | 0.397 (0.198 to 0.601) | 0.441 (0.201 to 0.656) | 0.608 (0.312 to 0.864) | 0.476 (0.231 to 0.704) |
| **NRI-** | -0.069 (-0.079 to -0.061) | -0.069 (-0.075 to -0.062) | -0.106 (-0.110 to -0.102) | -0.111 (-0.115 to -0.108) | -0.119 (-0.127 to -0.111) | -0.113 (-0.118 to -0.107) |
| **Continuous NRI** |  |  |  |  |  |  |
| **NRI** | 0.199 (0.116 to 0.283) | 0.214 (0.161 to 0.275) | 0.155 (-0.211 to 0.478) | 0.366 (0.001 to 0.701) | 0.077 (-0.238 to 0.324) | 0.122 (-0.204 to 0.455) |
| **NRI+** | 0.328 (0.252 to 0.402) | 0.324 (0.274 to 0.382) | 0.450 (0.084 to 0.770) | 0.480 (0.117 to 0.816) | 0.748 (0.435 to 0.999) | 0.594 (0.256 to 0.928) |
| **NRI-** | -0.129 (-0.145 to -0.112) | -0.110 (-0.121 to -0.097) | -0.295 (-0.305 to -0.284) | -0.114 (-0.124 to -0.104) | -0.672 (-0.688 to -0.655) | -0.472 (-0.488 to -0.457) |

**eTable 11. Performance Measures of the AI-ECG Model Output Probability, Pooled Cohort Equations to Prevent Heart Failure, and Predicting Risk of Cardiovascular Disease Events Equations for Predicting Incident Heart Failure with Censoring the Observations at the Median Duration of Follow-up.** Abbreviations: ELSA-Brasil, Brazilian Longitudinal Study of Adult Health; NPV, negative predictive value; PCP-HF, Pooled Cohort Equations to Prevent Heart Failure; PPV, positive predictive value; PREVENT, Predicting Risk of Cardiovascular Disease Events; UKB, UK Biobank; YNHHS, Yale New Haven Health System.

| **Metric** | **YNHHS** | | | **UKB** | | | **ELSA-Brasil** | | |
| --- | --- | --- | --- | --- | --- | --- | --- | --- | --- |
|  | **PCP-HF** | **PREVENT** | **AI-ECG** | **PCP-HF** | **PREVENT** | **AI-ECG** | **PCP-HF** | **PREVENT** | **AI-ECG** |
| **Sensitivity** | 47.4% (43.0-51.7) | 46.3% (43.6-49.1) | 59.9% (57.1-62.6) | 15.1% (1.2-29.1) | 6.7% (0-15.6) | 50.6% (31.3-69.9) | 12.5% (0-25.8) | 18.5% (5.0-32.1) | 65.5% (48.0-83.0) |
| **Specificity** | 68.0% (66.9-69.1) | 68.5% (67.8-69.2) | 73.8% (73.1-74.5) | 97.6% (97.4-97.9) | 98.2% (98-98.4) | 89.4% (88.9-89.8) | 97.9% (97.5-98.3) | 97.6% (97.2-98) | 86.7% (85.9-87.5) |
| **PPV** | 7.0% (6.1-7.9)% | 7.2% (6.6-7.8) | 10.8% (10.0-11.5) | 0.6% (0-1.3) | 0.4% (0-0.8) | 0.4% (0.2-0.7) | 1.6% (0-3.5) | 1.9% (0.4-3.3) | 1.2% (0.7-1.7) |
| **NPV** | 96.2% (95.8-96.7) | 96.0% (95.7-96.3) | 97.2% (97.0-97.4) | 99.9% (99.9-99.9) | 99.9% (99.9-99.9) | 99.9% (99.9-100) | 99.8% (99.6-99.9) | 99.8% (99.7-99.9) | 99.9% (99.8-100) |

**eTable 12. Age- and Sex-Adjusted Cox Proportional Hazard Models for the Prediction of Non-Heart Failure Clinical Outcomes.** Abbreviations: AMI, acute myocardial infarction; ELSA Brasil, Brazilian Longitudinal Study of Adult Health; HF, heart failure; UKB, UK Biobank; YNHHS, Yale New Haven Health System

| **Outcome** | **YNHHS** | **UKB** | **ELSA-Brasil** |
| --- | --- | --- | --- |
| **Primary AMI Hospitalization** | 1.12 (0.89-1.40) | 1.23 (0.85-1.76) | 3.00 (1.78-5.08) |
| **Primary Stroke Hospitalization** | 1.18 (1.09-1.27) | 1.54 (1.10-2.15) | 3.86 (2.28-6.51) |
| **All-cause Death** | 0.99 (0.95-1.03) | 1.58 (1.22-2.05) | 2.18 (1.65-2.88) |
| **Major Adverse Cardiovascular Events** | 1.76 (1.70-1.82) | 1.62 (1.37-1.93) | 2.68 (2.14-3.36) |

**eTable 13. Age- and Sex-Adjusted Cox Proportional Hazard Models for the Prediction of Non-Heart Failure Clinical Outcomes Across Yale New Haven Health System Sites.** Abbreviations: AMI, acute myocardial infarction; HF, heart failure; NEMG, Northeast Medical Group; L&M, Lawrence and Memorial Hospital; YNHH, Yale New Haven Hospital

| **Outcome** | **YNHH** | **Bridgeport** | **Greenwich** | **L&M** | **Westerly** | **NEMG** |
| --- | --- | --- | --- | --- | --- | --- |
| **Primary AMI Hospitalization** | 1.33 (0.73-2.42) | 1.31 (0.70-2.44) | 2.05 (0.71-5.88) | 1.68 (0.85-3.29) | 1.93 (0.82-4.55) | 0.76 (0.55-1.05) |
| **Primary Stroke Hospitalization** | 1.11 (0.98-1.26) | 1.20 (0.99-1.45) | 1.34 (1.07-1.67) | 1.24 (1.04-1.49) | 1.12 (0.81-1.55) | 1.05 (0.86-1.28) |
| **All-cause Death** | 0.95 (0.90-1.01) | 1.01 (0.91-1.12) | 1.24 (1.08-1.43) | 0.99 (0.88-1.12) | 0.95 (0.77-1.17) | 1.00 (0.90-1.11) |
| **Major Adverse Cardiovascular Events** | 1.73 (1.65-1.81) | 1.66 (1.53-1.80) | 1.77 (1.58-1.98) | 1.93 (1.76-2.12) | 1.57 (1.35-1.83) | 1.70 (1.56-1.85) |
